# Metabolite-based dietary supplementation in human type 1 diabetes is associated with microbiota and immune modulation

**DOI:** 10.1101/2021.09.15.21263059

**Authors:** Kirstine J. Bell, Sonia Saad, Bree J. Tillett, Helen M. McGuire, Sara Bordbar, Yu Anne Yap, Long T. Nguyen, Marc R. Wilkins, Susan Corley, Shannon Brodie, Sussan Duong, Courtney J. Wright, Stephen Twigg, Barbara Fazekas de St Groth, Leonard C. Harrison, Charles R. Mackay, Esteban N. Gurzov, Emma E. Hamilton-Williams, Eliana Mariño

**Affiliations:** Charles Perkins Centre, University of Sydney, Camperdown, NSW 2050 Australia; Faculty of Medicine and Health, Sydney Medical School, University of Sydney, NSW Australia; Kolling Institute of Medical Research, Royal North Shore Hospital, Sydney Medical School, University of Sydney, St Leonards, NSW, Australia; The University of Queensland Diamantina Institute, The University of Queensland, Woolloongabba, QLD 4102, Australia; Discipline of Pathology, Faculty of Medicine and Health, The University of Sydney, Camperdown, NSW 2050 Australia; Ramaciotti Facility for Human Systems Biology, The University of Sydney, Camperdown, NSW 2050 Australia; Infection and Immunity Program, Biomedicine Discovery Institute, Department of Biochemistry, Monash University, Melbourne, Victoria 3800, Australia; Systems Biology Initiative, School of Biotechnology and Biomolecular Sciences, University of New South Wales, NSW, 2052, Australia; Walter and Eliza Hall Institute of Medical Research, Parkville, Victoria 3052, Australia and Department of Medical Biology, University of Melbourne, Victoria 3010, Australia; Signal Transduction and Metabolism Laboratory, Université libre de Bruxelles, Brussels 1070, Belgium

**Keywords:** Type 1 diabetes, SCFAs, microbiota, immune regulation, dietary-metabolites, autoimmunity.

## Abstract

**Background:** Short-chain fatty acids (SCFAs) produced by the gut microbiota have beneficial anti-inflammatory and gut homeostasis effects and prevent type 1 diabetes (T1D) in mice. Reduced SCFA production indicates a loss of beneficial bacteria, commonly associated with chronic autoimmune and inflammatory diseases, including T1D and type 2 diabetes. Here we addressed whether a metabolite-based dietary supplement has an impact on humans with T1D. We conducted a single-arm pilot-and-feasibility trial with high-amylose maize resistant starch modified with acetate and butyrate (HAMSAB) to assess safety, while monitoring changes in the microbiota in alignment with modulation of the immune system status.

**Results:** HAMSAB supplement was administered for six weeks with follow-up at 12 weeks in adults with long-standing T1D. Increased concentrations of SCFA acetate, propionate, and butyrate in stools and plasma were in concert with a shift in the composition and function of the gut microbiota. While glucose control and insulin requirements did not change, subjects with the highest SCFA concentrations exhibited the best glycemic control. *Bifidobacterium longum*, *Bifidobacterium adolescentis*, and vitamin B7 production correlated with lower HbA1c and basal insulin requirements. Circulating B and T cells developed a more regulatory phenotype post-intervention.

**Conclusion:** Changes in gut microbiota composition, function, and immune profile following six weeks of HAMSAB supplementation were associated with increased SCFAs in stools and plasma. The persistence of these effects suggests that targeting dietary SCFAs may be a mechanism to alter immune profiles, promote immune tolerance and improve glycemic control for the treatment of T1D.

**Trial registration:** ACTRN12618001391268. Registered 20 August 2018, https://www.anzctr.org.au/Trial/Registration/TrialReview.aspx?id=375792

## Background

Short chain fatty acids (SCFAs) are metabolites produced by the gut microbiota, that greatly affect human health and disease [1]. Mostly produced from fermentation of non-digestible dietary carbohydrates, SCFAs, primarily acetate, propionate, and butyrate are anti-inflammatory and critical for maintaining gut homeostasis as well as playing a role in host energy metabolism [2]. Reduced SCFA production is an indicator of a loss of beneficial bacteria (dysbiosis), which is commonly associated with chronic autoimmune and inflammatory diseases including type 1 diabetes (T1D) and type 2 diabetes [3, 4]. Indeed, studies in mice and humans have shown that a deficiency in production of SCFAs by the gut microbiota is associated with the development of T1D, starting well before clinical diagnosis [5–8], which may be linked to altered gut homeostasis [9–11]. Insufficient intake of dietary fiber, or a gut microbiota that is poor at SCFA production may underpin the increased incidence of T1D and many other Western diseases [1, 2]. Thus, a microbiota-targeted dietary intervention that tackles the underlying functional dysbiosis (i.e. deficiency of SCFAs and altered microbiota function) may have great potential in humans to prevent or treat T1D, as it does in autoimmune diabetes-prone non-obese diabetic (NOD) mice [5].

Clinical studies have begun to show the potential of modulating the microbiota composition via the use of prebiotics, probiotics and fecal transplantation as an alternative approach to treat inflammatory diseases [12]. Most microbiota-based therapies identify bacterial communities or pathways associated with disease and then aim to restore specific health-associated species, an approach with inherent caveats. For example, the wide inter-individual variability in the gut microbiota associated with diseases such as T1D [3, 7] and key factors such as diet and host genetics, which affect microbiota colonization and adaptation, are all barriers to current investigative approaches [13]. Post-biotic targets (metabolites produced by the gut microbiota) have recently emerged as a novel alternative to promote health and circumvent these barriers [14]. We have developed a high-SCFA-yielding dietary supplements that ameliorates gut infection, improves chemotherapy efficacy and protects mice against T1D via regulating the immune system [5, 10, 15]. The SCFA-based supplement is a type 2 resistant starch consisting of a high amylose (70%) maize starch (HAMS) that has been modified by bonding the acetate and butyrate (HAMSAB). It is resistant to digestion in the upper gastrointestinal tract, delivers a very high yield of SCFAs in the colon, and is a powerful tool to assess the effects of SCFAs on intestinal biology [5, 10, 15, 16]. The HAMSAB diet prevented beta-cell destruction by T cells and protected against T1D in 90% of NOD mice. Protection against T1D was associated with microbiota composition changes. The mechanism behind this SCFA-induced T1D protection involved synergistic effects of acetate and butyrate; expansion of regulatory T cells (Treg) was butyrate-dependent and a decrease in pathogenic B cells, CD4^+^ and CD8^+^ T cells was acetate-dependent [5]. Therefore, this dietary intervention targets microbiota-host interactions, nutrition, and phylogeny.

Several human studies support the positive effects of increasing total fiber consumption in the form of high-amylose maize resistant starch (HAMS) or butyrate-yielding HAMS (HAMSB) supplementation alone in glycemic control [17, 18] or gastrointestinal conditions [19–21]. We report the first interventional study in humans with T1D to determine the effects of delivering a combination of dietary acetate and butyrate on the immune system via the gut microbiota. We therefore, conducted a single-arm pilot-and-feasibility study using HAMSAB administered for 6 weeks with follow-up at 12 weeks in adults with long-standing T1D.

## Results

### Clinical characteristics and protocol adherence

A total of 25 subjects between 18-45 years of age, diagnosed with T1D for at least 6 months and had HbA1c ≤ 8.5% were screened. Among this group, 20 subjects commenced HAMSAB supplementation (Fig. 1). Participants consumed 40g/day of the HAMSAB supplement (20g in the morning and at night) for 6 weeks with follow-up at 12 weeks. Participants had been diagnosed with T1D for a mean of 14 ± 12 years, 60% used multiple daily injection insulin therapy and mean HbA1c at baseline was 7.1 ± 0.6% (54 ± 7 mmol/mol) (Table 1). Two subjects withdrew, and one needed to halve the dose from week 3 onwards due to gastrointestinal side effects, as per protocol. Eighteen (90%) of 20 subjects starting the HAMSAB supplement remained in the trial and on the full dose for the entire six weeks (Fig. 1). Participants consumed 94 ± 10% of all doses based on the participants self-reported logbooks and 95 ± 8% based on returned unused supplement. Thus, indicating high compliance with the intervention. Mean HbA1c remained unchanged from baseline (W0) at both week 6 (W6) and week 12 (W12) (Table 1). Based on the continuous glucose monitoring (CGM) data, the mean daily average blood glucose and the mean standard deviation of CGM blood glucose values were not significantly different across the study (Table 1). Insulin dosing also remained stable throughout the study with no difference in the proportion of basal versus bolus insulin (Table 1). Only 31.5% (6/19) participants had detectable fasting plasma C-peptide measured by multiplex at the start of the study, the concentration of which did not change (data not shown). However, plasma C-peptide was not detected at fasting or at any timepoint during the 2-hour Mixed Meal Tolerance Test (MMTT; data not shown).

**Figure. 1.**
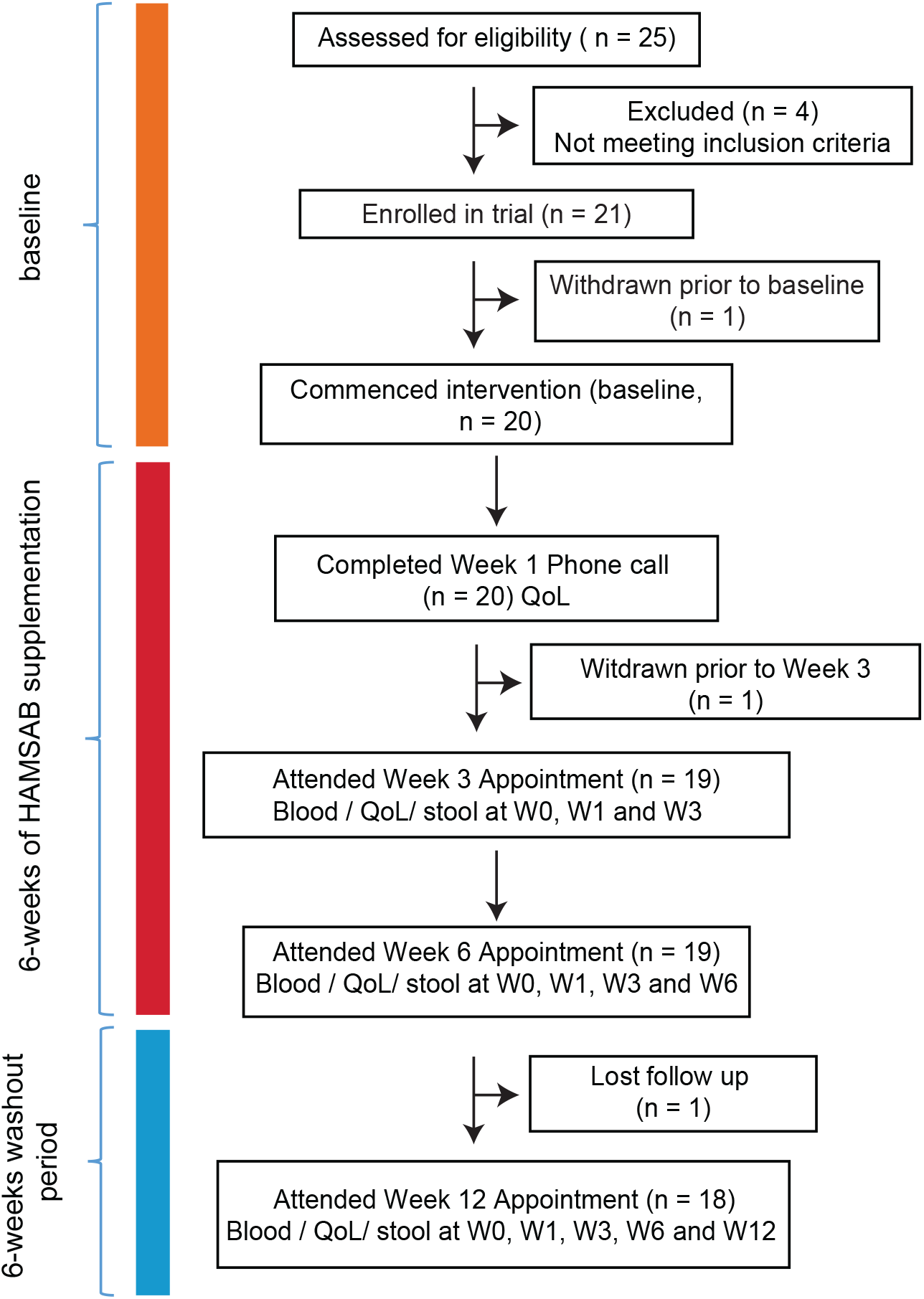
Experimental study in patients with T1D. Schematic diagram illustrating the study design and selection procedure for the individuals enrolled in the study.

**Table 1.**
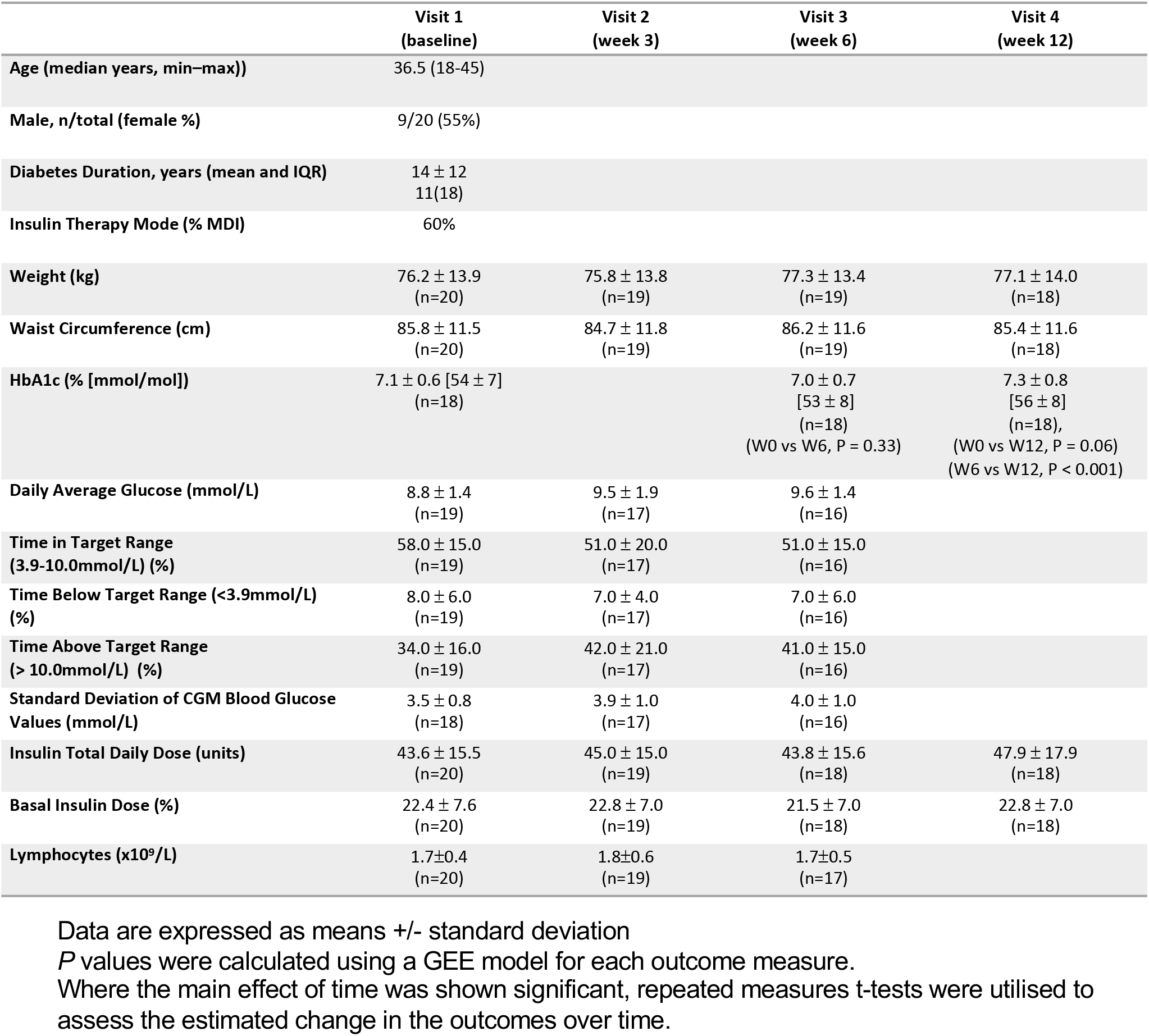
Baseline and follow-up characteristics. Data are expressed as means +/-standard deviation. P values were calculated using a GEE model for each outcome measure. Where the main effect of time was shown significant, repeated measures t-tests were used to assess the estimated change in the outcomes over time.

|  | Visit 1<br>(baseline) | Visit 2<br>(week 3) | Visit 3<br>(week 6) | Visit 4<br>(week 12) |
| --- | --- | --- | --- | --- |
| Age (median years, min–max)) | 36.5 (18–45) |  |  |  |
| Male, n/total (female %) | 9/20 (55%) |  |  |  |
| Diabetes Duration, years (mean and IQR) | 14 ± 12<br>11(18) |  |  |  |
| Insulin Therapy Mode (% MDI) | 60% |  |  |  |
| Weight (kg) | 76.2 ± 13.9<br>(n=20) | 75.8 ± 13.8<br>(n=19) | 77.3 ± 13.4<br>(n=19) | 77.1 ± 14.0<br>(n=18) |
| Waist Circumference (cm) | 85.8 ± 11.5<br>(n=20) | 84.7 ± 11.8<br>(n=19) | 86.2 ± 11.6<br>(n=19) | 85.4 ± 11.6<br>(n=18) |
| HbA1c (% [mmol/mol]) | 7.1 ± 0.6 [54 ± 7]<br>(n=18) |  | 7.0 ± 0.7<br>[53 ± 8]<br>(n=18)<br>(W0 vs W6, P = 0.33) | 7.3 ± 0.8<br>[56 ± 8]<br>(n=18),<br>(W0 vs W12, P = 0.06)<br>(W6 vs W12, P < 0.001) |
| Daily Average Glucose (mmol/L) | 8.8 ± 1.4<br>(n=19) | 9.5 ± 1.9<br>(n=17) | 9.6 ± 1.4<br>(n=16) |  |
| Time in Target Range<br>(3.9–10.0mmol/L) (%) | 58.0 ± 15.0<br>(n=19) | 51.0 ± 20.0<br>(n=17) | 51.0 ± 15.0<br>(n=16) |  |
| Time Below Target Range (<3.9mmol/L)<br>(%) | 8.0 ± 6.0<br>(n=19) | 7.0 ± 4.0<br>(n=17) | 7.0 ± 6.0<br>(n=16) |  |
| Time Above Target Range<br>(> 10.0mmol/L) (%) | 34.0 ± 16.0<br>(n=19) | 42.0 ± 21.0<br>(n=17) | 41.0 ± 15.0<br>(n=16) |  |
| Standard Deviation of CGM Blood Glucose<br>Values (mmol/L) | 3.5 ± 0.8<br>(n=18) | 3.9 ± 1.0<br>(n=17) | 4.0 ± 1.0<br>(n=16) |  |
| Insulin Total Daily Dose (units) | 43.6 ± 15.5<br>(n=20) | 45.0 ± 15.0<br>(n=19) | 43.8 ± 15.6<br>(n=18) | 47.9 ± 17.9<br>(n=18) |
| Basal Insulin Dose (%) | 22.4 ± 7.6<br>(n=20) | 22.8 ± 7.0<br>(n=19) | 21.5 ± 7.0<br>(n=18) | 22.8 ± 7.0<br>(n=18) |
| Lymphocytes (x10<sup>9</sup>/L) | 1.7±0.4<br>(n=20) | 1.8±0.6<br>(n=19) | 1.7±0.5<br>(n=17) |  |

Data are expressed as means +/- standard deviation P values were calculated using a GEE model for each outcome measure. Where the main effect of time was shown significant, repeated measures t-tests were utilised to assess the estimated change in the outcomes over time.

Gastrointestinal symptoms in relation to HAMSAB intake remained consistent or improved throughout the trial (Table S1A). Significantly higher symptom scores, indicating greater quality of life, were reported in W1 and W12 compared to baseline (Table S1A). Consistent with our data, other resistant starch intervention trials have reported that participants with no functional gastrointestinal disorders can tolerate high daily doses of resistant starch up to 50g per day [18, 22–24]. No changes in nutritional intake, assessed by 3-day food diaries, were observed between W0 and W6 (Table S1B). Weight and waist circumference remained stable throughout the trial (Table 1). Nineteen participants completed the supplement acceptability survey at W3 and W6. The majority of participants found the supplement acceptable through the length of the treatment. Twenty-six adverse events were reported during the trial, but approximately half (12/26) were unrelated to either the supplement or study protocol. The remainder related to minor gastrointestinal side-effects. No serious adverse events and no toxicity events were associated with the HAMSAB supplement dose or time on supplementation. Thus, data showed a dose of 40g of HAMSAB supplement was safe and tolerable in adults with T1D.

### HAMSAB supplementation is followed by increased SCFAs in stool and circulation

Similar to mice, several studies have reported that humans with T1D have reduced concentrations of stool and plasma SCFAs [5, 6, 8]. Consumption of the HAMSAB supplement over six weeks was followed by increased acetate, butyrate and propionate concentrations in both stool and plasma (Fig. 2). In the stool, the majority (77.7%) of subjects had >2-fold increase in acetate and propionate, and 61% subjects had >2-fold increase in butyrate concentration at W6 (Fig. 2A). Following the 6-week washout period, over 50% of subjects continued to have >2-fold higher concentration of SCFAs in the stool compared to baseline. In plasma, acetate concentrations started to rise significantly after three weeks on HAMSAB supplementation reaching a maximum peak at W6 (58% subjects >2-fold plasma acetate increased, Fig. 2B). To a lesser but also significant degree, propionate and butyrate increased in the plasma at W6. At W12, at the completion of the 6-week washout period, 8/19 subjects continued to maintain elevated concentrations of acetate in the plasma, but not propionate or butyrate. Thus, an increased SCFA availability in both the large intestine and circulation occurred after HAMSAB supplementation.

**Figure. 2.**
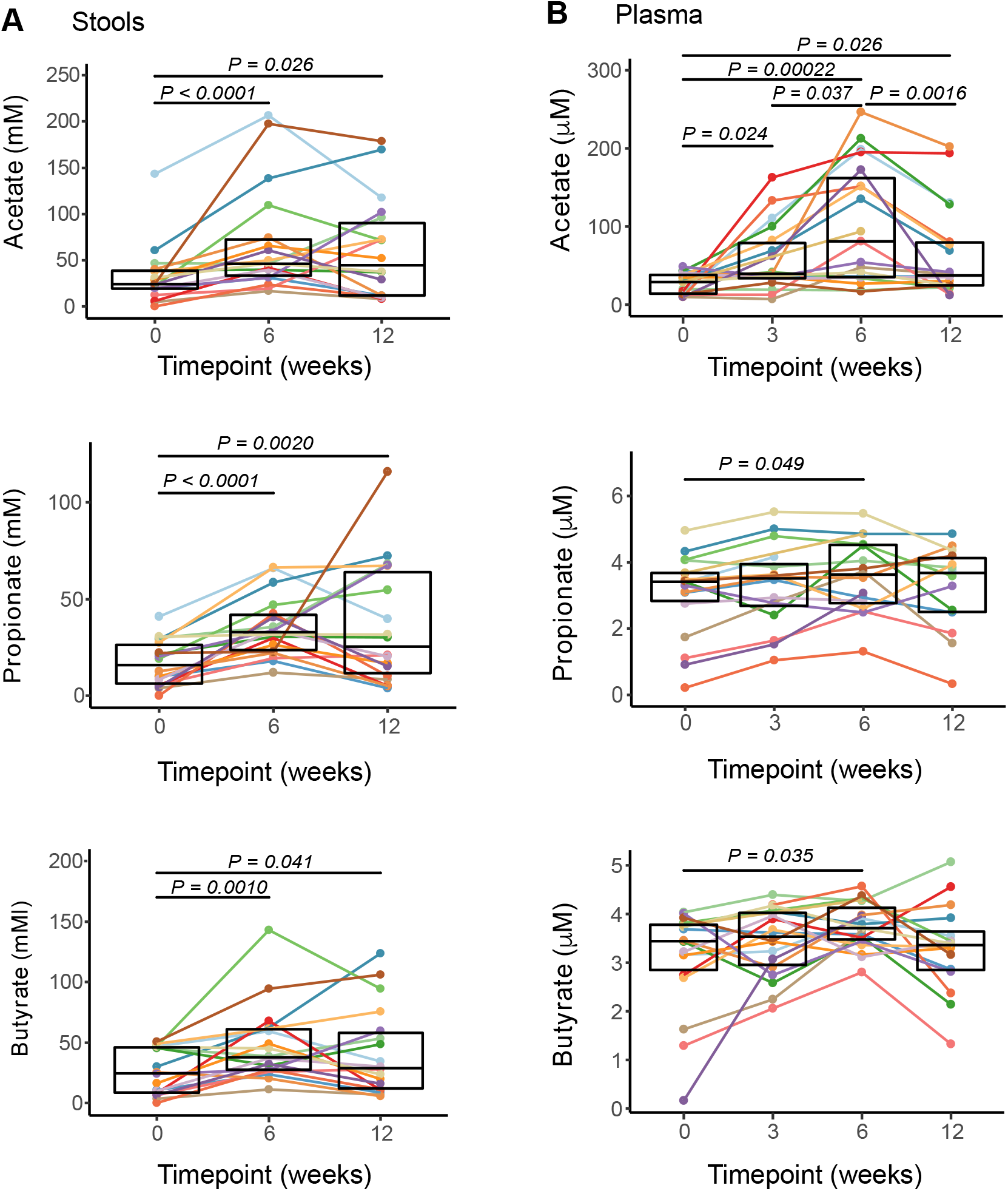
Increased concentration of short chain fatty acids in stool and plasma following HAMSAB supplementation. (A) Acetate, propionate and butyrate concentrations in stool (mM) and (B) plasma (µM). Overall significance determined by GEE and pairwise differences between timepoints by estimated marginal means and include a Tukey adjustment for multiple corrections. Colours indicate individual subjects. Box plots show mean and upper and lower quartile ranges.

### HAMSAB delivery is linked with changes in the gut microbiota

High-depth metagenomic sequencing of the stool microbiota found that taxonomic composition of the microbiome at W6 differed from baseline and W12, which were more similar in composition (Fig. 3A). Multivariate sPLS-DA analyses determined the changes in the microbiome at W6 were associated with unclassified members of the genus *Parabacteroides, Parabacteroides distasonis*, *Parabacteroides merdae* as well as *Bacteroides ovatus, Bifidobacterium adolescentis and Dialister invisus* (Fig. 3A and Fig. 1SA). In line with this change, alpha diversity was reduced at W6 on completion of the supplement, returning to similar diversity as baseline at W12 (Fig. 3B). Univariate analyses also identified significant increases in *Bacteroides uniformis,* unclassified *Parabacteroides* and *P. distasonis* from baseline to W6 (Fig. 3C). Meanwhile, *Eubacterium ramulus, Eubacterium eligens* and *Coprococcus comes*, which are known to feed on other carbohydrate sources such as pectin and flavonoids [25–27], were all decreased from baseline to W6 (Fig. 3C). *B. adolescentis* and *B. ovatus* marginally increased at W6 and then dropped back at W12. *Alistipes putredinis* showed the opposite trend, marginally decreasing at W6 and then significantly increasing at W12. Together, these data show a profound change in the composition of the gut microbiota following the HAMSAB diet, likely indicating expansion of bacteria able to use the delivered supplement.

**Figure 3.**
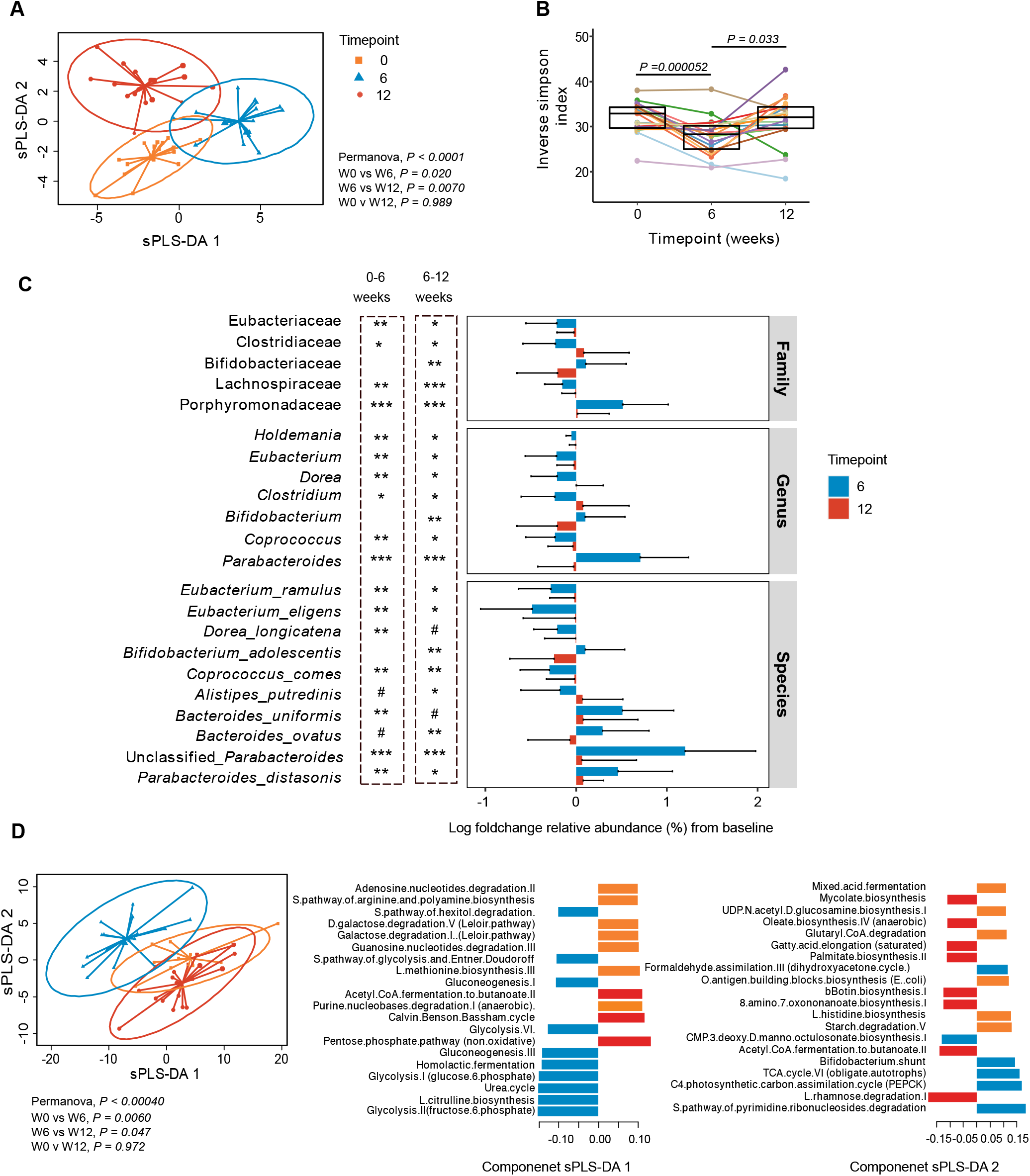
Composition and function of the gut microbiome is altered following HAMSAB supplementation. (A) Multivariate sPLS-DA comparing microbial species present at each timepoint. Significance determined by PERMANOVA. Loadings shown in Fig S1. (B) Alpha diversity measured by inverse Simpson index. Overall significance determined by GEE and pairwise differences between timepoints by estimated marginal means. Colours indicate individual participants. Box plots show mean and upper and lower quartile ranges. (C) Mean log foldchange in relative abundance from baseline at W6 and W12, grouped by taxonomic classification. Asterix represents GEE significance of changes in abundance from baseline. Error bars represent standard deviation. # adjusted *P* < 0.1, *adjusted *P* < 0.1-0.05, **adjusted *P* < 0.01, ***adjusted *P* < 0.001. (D) Multivariate sPLS-DA comparing microbial pathways present at each timepoint and plot loadings indicating the contribution of each bacterial function to the variance. Colour corresponds to the timepoint.

Next, a change in the metabolic pathways used by microbiota was investigated. The microbiome functions identified by metagenomic sequencing changed substantially at W6, but this was not sustained at follow-up (Fig. 3D). The pathways driving the changes at W6 included multiple carbohydrate energy production pathways including glycolysis and gluconeogenesis pathways, fermentation pathways such as homolactic fermentation and bifido-shunt, amino acid biosynthesis pathways (urea cycle and L citrulline biosynthesis) (Fig. 3D). Univariate comparisons identified 46 individual pathways that differed in abundance at W6 (Table S2). These were categorized within 15 superclasses. Glycolysis pathways, amino-acid biosynthesis pathways (urea cycle and L-citrulline biosynthesis) and the biosynthesis of cofactors and vitamins (B2 [flavin], B6 [pyridoxal], B7 [biotin] and B9 [folate]) all increased from baseline to W6, but this increase was not sustained at W12 follow-up. In contrast, Acetyl CoA fermentation to butanoate decreased at W6 and then increased again at W12, suggesting a switch occurs in SCFA production from starch utilization via glycolysis while on the HAMSAB supplementation, returning to SCFA production via fermentation at follow-up. Amino acid, nucleotide and vitamin biosynthesis can all be fueled by acetate utilization [10, 28]. These data indicate that HAMSAB supplementation was associated with a substantial shift in both the composition and function of the gut microbiota.

### HAMSAB supplementation result in increased circulating marginal zone B cells with a lower activation status

Previously, we reported that increasing SCFAs in NOD mice modulated autoimmune B and T cell responses in T1D and this was associated with protection against disease[5]. Thus, in a mechanistic analysis using mass-cytometry, we evaluated here whether HAMSAB supplementation changed immune cell subsets within peripheral blood mononuclear cells (PBMC). A multivariate PLS-DA visualization distinguished changes in the total proportion of multiple cell types across time (Fig. 4A). The overall changes in PBMCs were mainly seen at W12 compared to either baseline or W6 (Fig. 4A). Changes at W12 were related to cell subsets within the T cell and B cell compartments, along with the frequency of monocytes (Fig. 4B and gating strategy shown in Fig. S2). From this highly comprehensive approach covering more than 110 distinct immunophenotyping measurements, we identified 29 sub-populations that differed in frequency (% of live) and 17 that differed in the mean signal intensity (MSI) of phenotypic markers (Fig. S3A, B). The frequency of total B cells was increased at W6 of HAMSAB supplementation, and remained elevated after the 6-week washout (Fig. 4C and Fig. S3A). The increased frequency of B cells at W6 was mainly the result of an increase in naïve B cells identified as IgD^+^CD27^-^ (Fig. 4C and Fig. S3A). After the 6-week washout period, CD86 expression was downregulated on B cells gated as CD27^+^IgD^+^ IgM^hi^ (Fig. 4D and Fig. S3B). This CD27^+^IgD^+^IgM^hi^ subpopulation of human B cells has been described as phenotypically related to marginal zone B (MZB) cells [29, 30]. Downregulated CD86 expression on MZB cells has also been observed in the spleen and pancreatic lymph nodes of HAMSA-fed NOD mice in which reduction of hyperactive antigen-presenting MZB cells correlated with protection against diabetes [5, 31]. After termination of the HAMSAB supplementation, we observed an increase in the frequency of atypical IgD^-^IgM^-^ B cells (Fig. 4E), which have been detected in the blood of patients with systemic lupus erythematosus (SLE) and have defective gut-associated MZB development [32].

**Figure 4.**
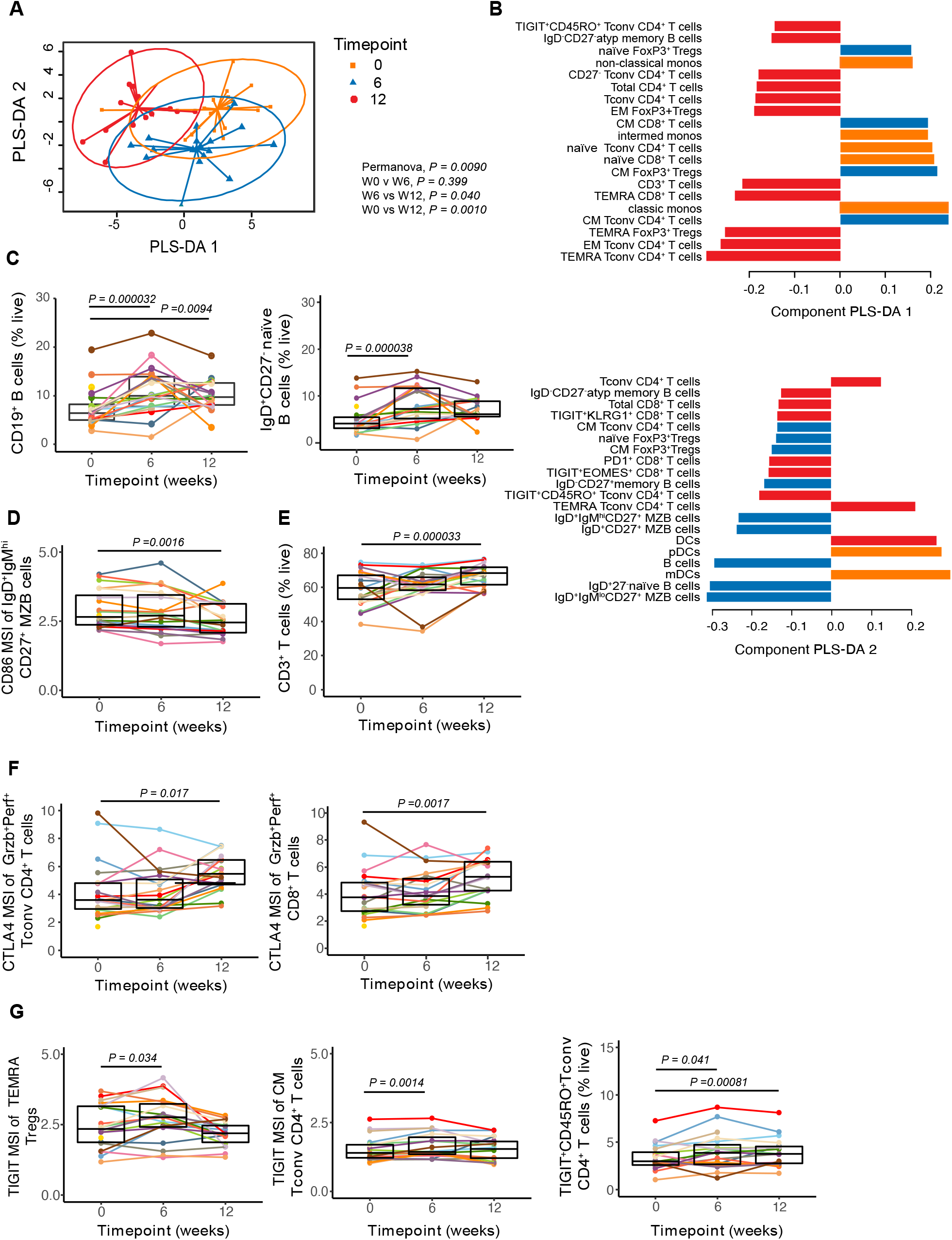
HAMSAB supplementation is accompanied by modulation of the immune system at W6 and W12 follow-up. (A) Multivariate PLS-DA comparing the proportions of major immune populations assessed by mass cytometry within total live cells. Significance determined by PERMANOVA. (B) PLS-DA plot loadings indicating the contribution of each immune population. (C) Proportions of total CD19^+^ B cells and IgD^+^CD27^-^ naïve B cells within live cells. (D) IgD^+^IgM^hi^CD27^+^ MZ B cells expressing CD86 (mean geometric signal intensity, MSI). (E) CD3^+^ T cell % within live cells. (F) CTLA4 expression (MSI) on granzyme B^+^ perforin^+^ (Grzb^+^Perf^+^) Tconv CD4^+^ T cells and Grzb^+^Perf^+^ CD8^+^ T cells. (G) TIGIT expression (MSI) on TEMRA Tregs, CM Tconv CD4^+^ T cells and % TIGIT^+^CD45RO^+^ Tconv within live cells. Coloured dots and lines represent each subject. Box plots show mean and upper and lower quartile ranges. Significance determined by GEE. Adjusted *P* values are (6W vs W0) or (12W vs W0). Gating strategy shown in Fig. S2.

### Increased immunoregulatory changes within T cells following HAMSAB supplementation

The total frequency of CD3^+^ T cells increased after HAMSAB supplementation, reaching a peak at W12 (Fig. 4E and Fig. S3A). This was reflected by an increase in effector memory (EM) and terminal effector memory CD45RA^+^ (TEMRA) cells within both CD4^+^ and CD8^+^ T cell compartments (Figs. 3A, 4A-D). EM and TEMRA were increased in both CD4^+^ regulatory T cells (Tregs) and non-regulatory conventional CD4^+^ T cells (Tconv) (Figs. S3A, S4B). In contrast, the frequencies of naïve and central memory (CM) Tconv, Tregs and CD8^+^ T cells were significantly reduced at W12 compared with baseline (Figs. S3A, S4B-D). At W12, we observed a higher expression of CD152 (cytotoxic T-lymphocyte-associated protein 4-CTLA4) across EM and naïve Tconv and CD8^+^ T cell subsets (Figs. S3B and S4E). The fact that we found increased expression of inhibitory CTLA4 on Tconv and CD8^+^ T cells that expressed granzyme B and perforin (Grzb^+^Perf^+^) (Fig. 4F and Fig. S3B), suggests that HAMSAB may reprogram cytotoxic T cells. CTLA4 inhibits CD28-mediated T cell co-stimulation by binding and down-regulating CD80/86 on antigen-presenting cells [33], which is consistent with the lower CD86 expression on MZB cells, plasmacytoid dendritic cells (pDCs) and myeloid/conventional dendritic cells (mDCs) at W6 and W12 (Figs. S3B and S4F). The frequency of monocytes was decreased at W12 follow-up (Figs. S3A, S3B and S4G). We found that HAMSAB supplementation was followed by increased expression of the coinhibitory molecule TIGIT (T cell immunoreceptor with Ig and ITIM domains), which in Tregs has been shown to suppresses pro-inflammatory Th1 and Th17 responses [34] (Fig. 4G). TEMRA Foxp3^+^ Tregs and CM Tconv CD4^+^ T cells expressing TIGIT were found to be increased at W6, while CD45RO^+^ Tconv cells expressing TIGIT were increased at both W6 and W12. (Fig. 4G). Remarkably, our results show that the immune system in adults with long-standing T1D was modulated towards immune regulation following HAMSAB supplementation, which persisted after the treatment stopped.

### Circulating pro-inflammatory markers are decreased and oxidative phosphorylation is increased subsequent to HAMSAB supplementation

We quantified the plasma concentrations of several pro-inflammatory and anti-inflammatory cytokines by multiplex bead array (Fig. 5A and Fig. S5). Plasma IL-8, MIP-1α and bFGF were decreased following the 6-week washout period compared to baseline (Fig. 5A). IL-8 and MIP-1α are critical immune mediators associated with Th1 responses [35, 36]. The changes in immune cell types across the study suggested that the HAMSAB supplement might affect gene expression, given the potent metabolic-epigenetic effects of SCFAs reported previously [5, 37, 38]. RNA-seq from whole blood comparing baseline and W6, together with gene set analysis revealed that 52 KEGG pathways were upregulated (Table S3) including fatty acid metabolism (114 genes) and oxidative phosphorylation (31 genes), while no pathways were significantly downregulated at W6. Genes included in these pathways were not individually differentially expressed at FDR 0.05, consistent with our observation that only four genes overall showed a significant change at W6 (Table S4), but coordinated changes in pathways were evident (Fig. 5B, C and Table S3). Fatty acid metabolism has been suggested to play an important role in differentiation and regulation of T-cell function [39]. Likewise, resting lymphocytes produce energy via oxidative phosphorylation and fatty acid oxidation, and rapidly shift to aerobic glycolysis in activated lymphocytes [40]. These changes are in line with the immunoregulatory phenotype observed in the patients during and after HAMSAB supplementation. Together, these data again indicate that HAMSAB supplementation is associated with changes in immune profiles towards an immunoregulatory phenotype.

**Figure 5.**
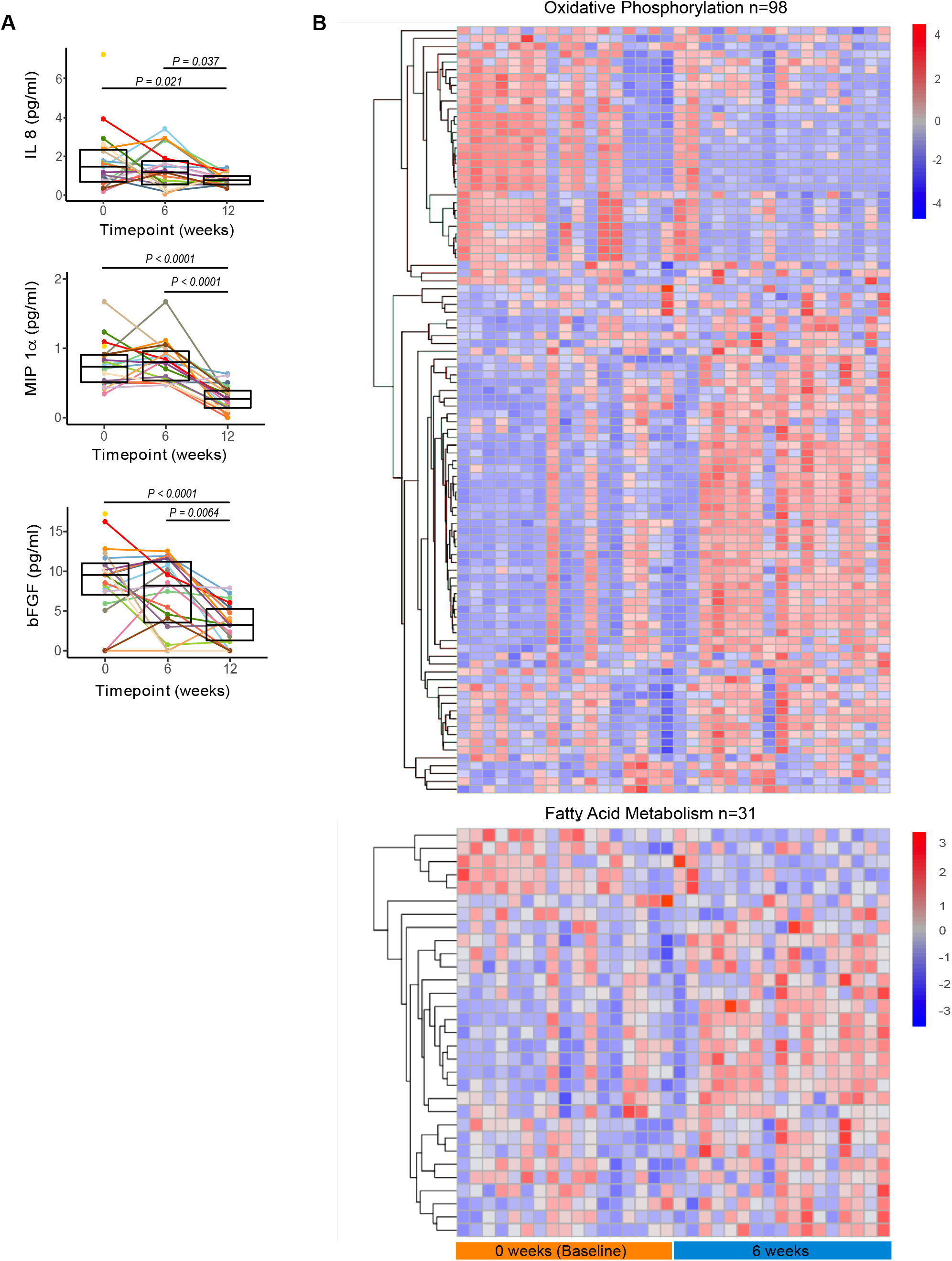
Circulating pro-inflammatory mediators are reduced at W12 in subjects following HAMSAB supplementation. (A) Serum IL-8, MIP1a and bFGF concentrations detected by multiplex assay. Overall significance determined by GEE and pairwise differences between timepoints by estimated marginal means. Box plots show mean and upper and lower quartile ranges. B) Hierarchical clustering of genes from fatty acid metabolism KEGG pathway gene set and from oxidative phosphorylation KEGG pathway gene set at baseline and 6-weeks of HAMSAB supplementation (FDR = 0.015 and 0.002, respectively).

### Evidence for interactions between microbiome, immune and clinical parameters

Next, we investigated the relationship between SCFAs, clinical parameters, metagenomic and immune phenotype data. At W6, plasma butyrate was negatively correlated with HbA1c, time below range and basal insulin dose (Fig. 6A, 6B and Fig. S6A, S6B), the later persisting beyond the 6-week washout period. Basal insulin dose and fecal acetate were also negatively correlated at W6 (Fig. S6A). There was no correlation between plasma acetate and glycemic variables (Fig. 6A) or between total daily insulin and SCFA concentrations at any time point (data not shown). Basal insulin dose negatively correlated with the relative abundance of *P. distasonis*, *B. longum, B. adolescentis* and *Streptococcus australis* and with the nucleoside/nucleotide degradation pathway pyrimidine riobonucleoside degradation across timepoints (Fig. 6C and Fig. S6C). HbA1c negatively correlated with *B. longum,* and biotin biosynthesis (Fig. 6C). These findings suggest that higher levels of *Bifidobacterium* taxa and biotin biosynthesis by the gut microbiota, supported by higher local acetate and butyrate availability, is associated with better glycemic control in adults with T1D.

**Figure 6.**
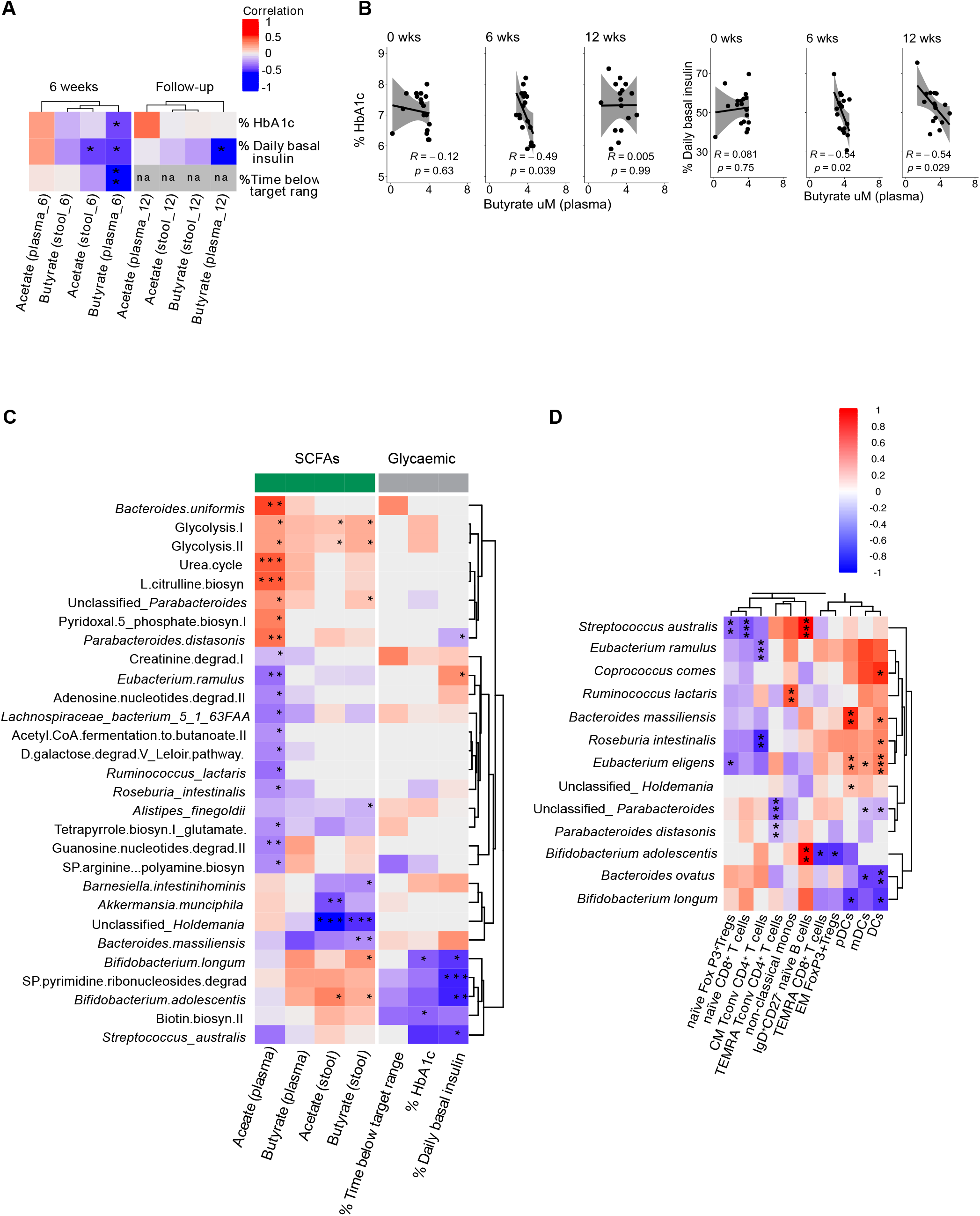
Increased SCFAs correlated with changes in glycemic control, commensal microbiota and immune cell changes. (A) Heatmap of Pearson r values between relative abundance of SCFAs and clinical data. * adjusted *P* < 0.05. ** adjusted *P* < 0.01. Stool and plasma short chain fatty acids are hierarchically clustered based on Bray-Curtis dissimilarity. (B) Pearson r values at each timepoint between plasma butyrate, HbA1c and daily basal insulin. Grey shading represents 95% confidence interval. (C) Heatmap of regression coefficients determined by GEEGLM between bacterial taxa and pathways with stool and plasma SCFAs and glycemic markers. Bacterial pathways and taxa are hierarchically clustered based on Bray-Curtis dissimilarity. (D) Heatmap of significant regression coefficients determined by GEEGLM between bacterial taxa that significantly changed across time (adj p<0.05) and/or those correlated with SCFA and glycemic markers in (C) and significantly altered immune subsets (adj p<0.05). * adjusted *P* < 0.05, ** adjusted *P* < 0.01, *** adjusted *P* < 0.001.

The increases in SCFAs in stool and plasma were related to changes in gut microbiota composition (Fig. 6C). Stool butyrate was positively correlated with unclassified *Parabacteroides, B. longum* and *B. adolescentis,* while stool acetate positively correlated only with *B. adolescentis.* Stool butyrate was negatively correlated with *Alistipes finegoldii, Barnesiella intestinohominis,* unclassified *Holdemania* and *Bacteroides massiliensis,* and stool acetate negatively correlated with *Akkermansia munciphila* and unclassified *Holdemania* (Fig. 6C). Likewise, plasma acetate was positively correlated with taxa that were most strongly increased at week 6 (*B. uniformis,* unclassified *Parabacteroides* and *P. distasonis*) and was negatively correlated with *E. ramulus*, *Lachnocpiraceae bacterium 5 1 63FAA*, *Ruminococcus lactaris* and *Roseburia intestinalis* (Fig. 6C). Although plasma butyrate showed similar trends to stool butyrate, these were not significant after correction for multiple comparisons (Fig. 6C). Plasma acetate correlated positively with several bacterial pathways: amino acid biosynthesis pathways (urea cycle and L-citrulline biosynthesis) and a B6 vitamin biosynthesis pathway (Fig. 6C). Glycolysis I and II pathways were positively correlated with stool acetate and butyrate and plasma acetate. Plasma acetate was negatively correlated with amine/polyamine biosynthesis, nucleoside/nucleotide degradation, fermentation and tetrapyrrole biosynthesis pathways (Fig. 6C). Hierarchical clustering of these correlations (Fig. 6C), clustered *B. longum, B. adolescentis*, *S. australis* and biotin biosynthesis away from the taxa and pathways correlated with plasma acetate. This indicates that a different microbial community may be associated with those subjects in whom a high level of acetate entered the circulation compared to those whose plasma acetate remained low.

Changes in immune subsets particularly those involving B cells, TIGIT^+^RO^+^ Tconv cells and non-classical monocytes were related to the increased concentrations of stool butyrate and plasma acetate (Fig. S6D). Further analysis of B cell subsets revealed that IgD^+^CD27^-^ naïve B cells positively correlated with *B. adolescentis* and *S. australis* (Fig. S6D). Naïve Tregs and naïve CD8^+^ T cells were negatively correlated with *S. australis.* Meanwhile, EM Tregs were negatively correlated with *B. adolescentis.* TEMRA Tconv cells were negatively correlated with *P. distasonis* and unclassified *Parabacteroides* (Fig. S6D). CM Tconv cells were negatively correlated with *E. ramulus* and *Roseburia intestinalis*. *E. eligens, Bacteroides massiliensis, Roseburia intestinalis* and *Coprococcus comes* were positively correlated with total DCs, mDCs and/or pDCs while *Bacteroides ovatus* and *B. longum* were negatively correlated with these subsets. In contrast, mDCs expressing CD86 were negatively correlated with *B.* ovatus (Fig. S6D and Fig. S4F). Overall, these data suggested that increased SCFA production following HAMSAB supplementation is linked with immune-microbiome interactions.

## Discussion

This is the first human study investigating the effects of an acetate- and butyrate-modified starch supplement (HAMSAB) in adults with long-standing T1D. Subjects taking the HAMSAB supplement increased acetate, propionate and butyrate in both stool and plasma, indicating metabolism of the starch by the gut bacteria. Changes in the gut bacterial composition and function included a reduction in carbohydrate degradation, promotion of gluconeogenesis, amino acid and vitamin B2, B6, B7 and B9 biosynthesis pathways, consistent with SCFA utilization by the microbiota. Remarkably, increased SCFAs following the HAMSAB supplement resulted in modulation of T cells, B cells, DCs and monocytes towards a more regulatory immune phenotype. A correlation network associated stool acetate and butyrate, plasma butyrate, higher *Bifidobacterium* and biotin biosynthesis, lower HbA1c and lower daily basal insulin requirements.

Subjects in our study consumed the HAMSAB supplement for 6 weeks, during which time they experienced no overall impacts on their diabetes management demonstrating the supplement was safe. While our study was not designed to determine efficacy, the observed correlations between increased SCFA and improved glycemic control were similar to what was found in human studies testing the effect of HAMS, type 4 resistant starches or high-fiber from vegetables in subjects with type 2 diabetes or metabolic disease, as well as in healthy individuals [4, 17, 41]. Provision of a high-fiber diet to adults with long-standing T1D for 6 months resulted in decreased daily blood glucose and decreased hypoglycemic events compared to a low-fiber diet [42]. An increased high-fiber diet in the form of oligofructose-enriched inulin, administered to children with long-standing T1D for 12-weeks, did not change HbA1c or inflammatory markers, but the inulin-treated children had preserved C-peptide and increased relative abundance of *Bifidobacterium* compared to the placebo group [43]. While our study could not assess whether the effects observed were due to the additional acetate and butyrate, preclinical mouse models in T1D, infection and cancer have shown the beneficial effects of added acetate or butyrate delivery compared to the starch (HAMS) alone [5, 10, 15]. Other studies have investigated the direct effect of oral postbiotics such as sodium butyrate changing the immune status in long-standing T1D and obese individuals [44, 45]. Postbiotic butyrate didn’t change NK cells, CD8^+^ T cells, Tregs or other immune cell subsets in individuals with long-standing T1D, but did reduce islet reactive IA-2-specific CD8^+^ T cells in butyrate treated individuals compared to placebo [44] and moderately affected cytokine responses in obese individuals [45]. As we observed reduced inflammatory markers and marked immune changes following supplementation, this suggests long-term HAMSAB dietary supplementation via the gut microbiota may elicit further beneficial effects from increased SCFAs over and above those from high-fiber or the starch carrier itself or from delivery of soluble SCFA alone.

HAMSAB supplementation was associated with significantly increased concentrations of SCFAs in stools and plasma. SCFAs are used as a source of energy by the host and the gut microbiota[46]. In humans, more than 95% of the bacterially produced SCFAs are absorbed by the colonocytes where butyrate is the major energy source [47–49]. Microbial SCFAs enter the blood circulation via the hepatic portal vein, where 70% and 30% of the acetate and propionate, respectively are taken up by the liver [50]. Acetate is used in the synthesis of cholesterol, long-chain fatty acids and amino acids such as glutamine and glutamate. Meanwhile, propionate acts as a precursor of glucogenesis [51–53]. Propionate and butyrate are mostly cleared in the liver before reaching portal circulation, which explains their lower concentrations in peripheral blood compared to stools. Released SCFAs that pass through the liver also are metabolized in many other organs such as the brain, heart, kidney, muscle, and adipose tissue where they intersect with their immune system and metabolism [1, 54]. Thus, the detection of SCFAs in peripheral blood from patients with T1D implies that these metabolites may have systemic functions, as shown in other human studies [55] [4, 56].

Increasing concentrations of acetate and butyrate after HAMSAB feeding were associated with protection against diabetes in NOD mice [5]. In our pre-clinical study, HAMSA diet led to a change in the overall composition of B cells, driven by a decrease in MZB cells with high CD86 and MHC-I expression. Meanwhile, the HAMSB diet drove an increase in regulatory T cell frequency and IL-10 production, and both diets reduced the proliferation of islet-specific T cells. Remarkably, these findings were similar to what we observed here, where MZB-like cells expressing CD86 decreased over time and CTLA4-expressing T cells increased. It has been shown that murine transitional two-marginal-zone B cells, which resemble murine IL-10-producing regulatory B cells (Bregs) and naïve human MZB cells, are induced by the gut microbiota to restrain chronic inflammation and protect against arthritis [57]. Moreover, acetate has been shown to induce the immunosuppressive activity of Bregs associated with reduced arthritis severity [58]. Hence, modulation of B cells towards a more regulatory phenotype observed in the patients at W6 of the HAMSAB delivery may have the potential to preserve pancreatic beta-cells if trialed in recent-onset or pre-clinical disease.

We did not investigate islet-specific T cells, as individuals with long-standing T1D have a low frequency of such cells [59, 60]. However, we observed an increase in TIGIT expressing cells similar to what has been shown with exhausted-like T cells associated with response to therapy in anti-CD3 and alefacept trials [61, 62]. Our results showing reduced frequency of monocytes, DCs and increased Treg-expressing CTLA-4 cells at W12 are consistent with previous reports that Treg-mediated CTLA4-dependent downregulation of CD80/CD86 on DCs could inhibit the rapid proliferation of pathogenic CD4^+^ T cells [63]. Data from the Abatacept trial administering the co-stimulation blocking CTLA-4-Ig in individuals with T1D showed that an increase in CM cell frequency correlated with a decline in beta-cell function [64, 65]. Consistent with this finding, the effects of HAMSAB supplementation were associated with a significant contraction in CM and naïve T cell populations. Likewise, it has been shown that an increase in monocytes is associated with the development of cardiovascular complications in long-standing T1D patients [66]. HAMSAB supplementation was associated with a reduction in the overall frequency of all three monocyte subsets. Changes in the monocyte subset balance, particularly an expansion in intermediate monocytes and an increase in CD45RO^+^ CD4^+^ T cells have been linked with beta-cell dysfunction in children with recent-onset T1D [67]. On the other hand, the changes we observed in classical monocytes after 6 weeks of HAMSAB supplementation are similar to what has been shown in healthy controls compared to patients with long-standing T1D and in recent-onset T1D patients within three months of diagnosis [68].

Changes in immune cell frequency were accompanied by a reduction in the inflammatory cytokines macrophage inflammatory protein 1-α (MIP-1α) and IL-8, which could indicate reduced innate immune cell activation. IL-8 is a neutrophil chemotactic factor, a reduction of which is likely to suggest amelioration of inflammation. IL-8 is associated with reduced levels of insulin growth factor-1 and related to poor metabolic control in adolescents with T1D [69]. Moreover, IL-8 is also produced from microbiota-induced NFκΒ activation pathways in colonocytes [70]. It has been shown in other probiotic diet trials that increased propionate leads to a reduction in IL-8 [71]. Similarly, MIP-1α is linked with the pathogenesis of various inflammatory diseases[72]. MIP-1α expression in the pancreas were associated with progression to T1D in NOD mice [73]. A relationship between bFGF and diabetes is unclear. Of note, the most significant changes in immune profile occurred at 12-weeks, when the levels of SCFAs in plasma and stools remained significantly elevated compared to the baseline, which may suggest delayed yet sustained effects of the supplement on immune system regulation. Further studies using a longer period of dietary intervention may be needed to assess optimal effects and benefits of the HAMSAB supplement.

Analysis of the fecal microbiome demonstrated a substantial shift in both the composition, diversity and function of the microbial community after the HAMSAB intervention. Other studies using specific resistant starches or dietary fiber have also shown decreases in the number of bacterial taxa due to differences in ability to metabolize resistant starch sources [24, 43, 74]. Major changes were specific to *P. distasonis,* which is a saccharolytic anaerobe involved in digestion of polysaccharides [75]. Two previous clinical trials that delivered HAMSB to healthy volunteers also observed increased abundance of *P. distasonis*, suggesting this is a common response to HAMS-modified starches [22, 23]. *P. distasonis* can produce acetate (though not a major producer) succinate and secondary bile acids [75, 76]. The health benefits of *P. distasonis* are variable and depend on the disease context. *P. distasonis* decreased weight gain, hyperglycemia and liver disease in a high-fat diet mouse model [76]. In a colitis model, *P. distasonis* components reduced disease severity [77]. In human disease, *Parabacteroides* genera have variously been found to be increased in children at the onset of T1D [78] and in ankylosing spondylitis [79]. As such, it is not yet clear whether an increase in *P. distasonis* abundance following HAMSAB treatment will have an overall beneficial impact in the context of T1D and further studies are needed to unravel the precise impact of this species, potentially using animal models.

The HAMSAB supplement promoted changes in *B. adolescentis*, which trended to increase while on the supplement, and was positively correlated with both acetate and butyrate concentration in the stool. Bifidobacteria do not produce butyrate themselves but are known for their mutualistic ‘cross-feeding’ behavior, whereby they produce lactate and acetate that support the presence of other butyrate-producing taxa [80]. Functional changes in the microbiota at 6-weeks included the ‘*bifidobacterium* shunt’, which is linked to this cross-feeding pathway. We observed that stool concentrations of acetate, butyrate and propionate all remained elevated at the 12-week follow up timepoint. This suggests that a persistent effect on SCFA availability may remain after the intervention stopped. Analysis of food-diaries and macronutrient intake suggested this was not due to any altered dietary patterns over the course of the study. Rather, it is likely that either SCFA production or utilization patterns changed. As we did not find any significant differences in the composition of the microbiota or the pathways present in the genomes of the bacteria at 12-weeks compared with baseline, this may represent a functional reprogramming of the taxa present or an alteration in the host consumption of SCFAs. A limitation of functional inference from metagenomic sequencing data is that it does not determine which pathways are actually active in the bacteria at any given time. Of note, the microbial pathway linked to fermentation to butanoate significantly increased at 12-weeks compared to 6-weeks, indicating a switch to SCFA production rather than utilization at 12-weeks. More direct measures of the host and bacterial activity are needed to clarify the mechanism, and such studies are ongoing.

The major microbial functional pathways that were altered at the 6-week timepoint included increased glycolysis/decreased carbohydrate degradation, increased amino acid synthesis and increased B group-vitamin and cofactor biosynthesis. The changes in carbohydrate metabolism suggest that the SCFAs released from HAMSAB were used as an energy source by the bacteria. *In vitro*, bacteria grown on acetate rather than glucose upregulate the TCA cycle and gluconeogenesis, consistent with our findings [10, 28]. The TCA cycle generates the precursor molecules that are required for the biosynthesis of nucleotides, amino acids, co-factors and vitamins. The biotin (vitamin B7) biosynthesis II pathway, which increased at week 6, clustered with *Bifidobacterium* and negatively correlated with HbA1c. *Bifidobacterium* do not possess biotin synthesis ability themselves, however; their establishment in the gut environment (particularly *B. longum)*, has been suggested to augment the production of the biotin precursor pymelate, which is metabolized into biotin by other species such as *Bacteroides uniformis* [81–83]. Biotin is an NFκB inhibitor with anti-inflammatory effects on the immune system, suppressing production of inflammatory cytokines [84].

Interestingly, features of the microbiota that correlated with improved glycemic control were linked to stool SCFAs and plasma butyrate concentrations but not with plasma acetate. However, only a subset of subjects increased acetate levels systemically, whereas almost all increased acetate in their stool. We speculate that this may be related to a difference in the host’s ability to absorb acetate, perhaps due to differences in intestinal permeability or differential utilization of acetate. Individuals with T1D have increased intestinal permeability compared to healthy subjects, although this characteristic is highly variable [85–87]. Furthermore, the microbiota associated with T1D are also highly variable, which may result in a different response to HAMSAB. A longer period of HAMSAB dietary intervention would help resolve these individual differences and potentially achieve greater effects of the HAMSAB supplement.

## Conclusion

The study was designed to determine the safety of the supplement in the context of T1D and it didn’t include a placebo control group. Although the explorative outcomes provided significant insights into the effects of HAMSAB supplementation on microbiota composition and immune status, the changes observed are associative from comparisons within participants over time. In addition, the intervention used was only 6 weeks, which is a relatively short timeframe to look at changes in markers such as HbA1c (usually looked at over 3-monthly intervals) or C-peptide (usually assessed over 12-month intervals). A follow-up placebo-controlled trial powered to investigate efficacy will be needed to determine casual effects from the HAMSAB supplement in a larger study and longer period of intervention to better determine any impact on glycemic control.

We have shown that HAMSAB supplementation remodels the gut microbiota and significantly impacts the immune system. Moreover, our study has validated our pre-clinical results in the NOD mouse, indicating that the HAMSAB supplement is a physiological and efficient way to modulate microbiota-host interactions in T1D. This trial provides the necessary data to allow for an appropriately powered, double-blinded, placebo-controlled randomized controlled trial in T1D. If efficacious, an anti-inflammatory dietary HAMSAB intervention should be a widely acceptable, non-invasive and highly accessible means of preserving beta-cell function in individuals with newly diagnosed T1D or in those at high risk of the disease.

## Methods

### Participants and study design

The trial was a single-arm pilot trial of the HAMSAB supplement administered twice daily as part of the usual diet for 6-weeks with follow-up at 12 weeks in adults with T1D. Inclusion criteria included: 18 to 45 years, males and females; clinical diagnosis of T1D for at least 6 months; HbA1c ≤ 8.5% and stable disease management. Exclusion criteria included: pregnancy or lactation, use of diabetes medications other than insulin, hypoglycaemia unawareness, concomitant disease or treatment that may impact on glycemic control, insulin requirements or other outcome measures, history or symptoms of gastrointestinal disease or malabsorption, any known conditions that could be associated with difficulty complying with the trial protocol, weight below 50kg or above 120kg, liver or renal disease, use of senor-augmented insulin therapy, unwillingness to maintain their normal stable diet during the study and/or following a restrictive diet that would impact their ability to take the supplement, recent (previous 6 weeks) or anticipated antibiotic use during the study.

Participants attended 4 visits at the Charles Perkins Centre Clinical Research Centre (W0, W3, W6 and W12). In the week prior to each study visit, participants wore a blinded continuous glucose monitor (Abbott Freestyle Libre Pro) for 14 days (up to W6). Optimal continual glucose monitoring (CGM) target in between 3.9-10 mmol/L. Participants completed: a 3-day food diary (2 weekdays and 1 weekend day via paper or smartphone app [Easy Diet Diary Research, Xyris], baseline and W6 only), a 7-day insulin logbook only if using multiple daily injections, otherwise insulin dosing history recorded from insulin pump, and the HAMSAB supplement dosing logbook (up to W6). Compliance with intervention was evaluated by self-reported log-books and measuring the returned unused supplement. Participants collected and froze a stool sample in the 24h prior to their appointment and completed three online questionnaires: a validated 18-item, 5-point likert scale Gastrointestinal Quality of Life Index Questionnaire[88]. Gastrointestinal symptoms (i.e. bloating, constipation and flatulence) measured as score range: 0-64, with higher scores indicating less gastrointestinal symptoms; a 10-item Diabetes-Specific Symptom Questionnaire (score range: 0-36 with higher scores indicating higher QoL) and an 8-item HAMSAB supplement feedback questionnaire to assess its palatability and acceptability. The HAMSAB supplement is a high amylose corn starch acetylated and butyrylated (Ingredion Incorporated, Bridgewater, NJ). To improve tolerability, participants commenced with 10g/day and increased the dose in 10g increments every 48h until 40g per day was reached. If the increased dose was not tolerated, the participant was to remain on the previous dose for an additional 48h before attempting the increased dose again. If that dose was not tolerated after the third attempt, (or on the judgement of the investigators), they were to remain on the previous dose for the remainder of the intervention. Participants were asked to return any unused HAMSAB supplement at the next visit.

### Laboratory measures

Blood and urine samples collected from participants at W0, W3 and W6 (n = 20, 19 and 19, respectively) were analyzed for serum urea, creatinine, electrolytes, liver function tests, bicarbonate, triglycerides and urinary albumin. Urinary pH analyzed by dipstick on-site by trial coordinator at time of collection. Food diaries were analyzed using nutrient analysis software (FoodWorks Professional 9, Xyris) to assess changes in food/nutrient intake. Continuous glucose monitoring data was included for all days where the monitoring was worn for a full 24h period (i.e. no partial days included). A Mixed Meal Tolerance Test (MMTT) was conducted at baseline, 6 and 12-weeks. Participants fasted for 8-12 hours and no bolus insulin dosed with the test drink or in the 3 hours prior. Usual basal insulin was allowed. The test was only performed if the fasting blood glucose was between 4-11mmol/L as described previously [89]. The test meal was a standard mixture of fat, carbohydrate and protein (Ensure; Nestle Health Care Nutrition, Inc). Venous blood samples were collected at 0, 30, 60, 90 and 120 minutes. Stimulated C-peptide was measured by ELISA (Sigma-Aldrich, RAB1389).

### SCFA analysis

Plasma samples were collected at baseline, W3, W6 and W12 (n = 20, 19, 19 and 18, respectively) and stool samples at baseline, W6 and W12 (n = 20, 19 and 18, respectively) and stored at −80°C until processing. SCFA acetate, propionate, and butyrate were measured in duplicates by gas chromatography after liquid-liquid extraction. Plasma SCFAs were directly measured by specialised polar phase gas chromatography-mass spectrometry (GC-MS) Phenomenex Zebron ZBFFAP column (Phenomenex, Torrance, California). Plasma (200μL) was mixed with internal standards (50µL of 200µM heptanoic acid internal standard and 50µL of 10 % sulfosalycilic acid). Samples were mixed with 30µL of 0.2M NaOH, centrifuged and let dry at room temperature. The residue was re-dissolved in 30μL of 1M phosphoric acid. Stool samples were mixed with internal standards solution and centrifuged. 100µL of supernatant was filtered and analyzed on an Agilent 7890A gas chromatograph (Agilent Technologies, Santa Clara, California). Peaks were detected with a flame ionization detector at 210°C and identified against calibration standards over the range 0 to 400mM.

### Mass cytometry

PBMC were isolated from blood collected at baseline, W6 and W12 (n = 20, 19 and 18, respectively) by Ficoll-paque (GE Healthcare, Uppsala) centrifugation, cryopreserved in 10% dimethyl sulfoxide, 20% foetal calf serum (FCS) and RMPI-1640 (Thermo Fisher Scientific, Waltham) and stored in vapour phase liquid nitrogen. All samples collected were thawed in batches, washed with RMPI-1640 (Thermo Fisher Scientific, Waltham), supplemented with 10% heat inactivated FCS and benzonase and subjected to immunophenotyping analysis by mass cytometry. A panel of 27 metal-tagged monoclonal antibodies supplied by the Ramaciotti Facility for Human Systems Biology was used for analysis (Extended Data Table 5). The gating strategy is shown in Extended Data Fig. 2. Unlabelled antibodies were purchased in a carrier-protein-free format and conjugated with the indicated metal isotope using the MaxPAR antibody conjugation kit (Fluidigm). For live-dead cell distinction, PBMCs were stained with 1.25µM cisplatin. Cells were incubated for 30 min initially with anti-CD45 antibodies conjugated to various metals, which facilitated barcoding and pooling of samples from multiple time points for each participant together prior to staining with the remaining antibodies targeting surface antigens. Cells were then fixed and permeabilised with FoxP3 staining kit (eBiosciences, San Diego) and stained with FoxP3 antibody. Cells were fixed in 4% paraformaldehyde containing DNA intercalator (0.125μM iridium-191/193; Fluidigm). After multiple washes, cells were diluted in MilliQ water containing 1:10 diluted EQ beads (Fluidigm) and acquired at a rate of 200–400 cells/second using a CyTOF 2 Helios upgraded mass cytometer (Fluidigm). All Helios data were normalised using the processing function within the CyTOF acquisition software based on the concurrently run EQ four element beads. Data analysis was performed using FlowJo version 10.4 software (FlowJo, LLC, Ashland, OR, USA). Samples were pre-gated on DNA^+^, live, CD45^+^ cells, and exported for further analysis.

### Metagenomic sequencing and data processing

Stool samples from participants were collected at baseline, W6 and W12 (n = 20, 19 and 18, respectively) and DNA was extracted from 0.2g stool using a repeated bead beating method [90]. Briefly, bacteria were lysed and homogenised by bead beating (0.5 mm zirconia beads) with lysis buffer containing 4% SDS, 500nM NaCl, 60mM Tris-HCL and 50mM EDTA using a Precellys24 homogenizer (Bertin Technologies) for 3x 60sec cycles at 5000 rpm with 15sec rests. Bacterial lysis was enhanced by further incubating at 70^°^C for 15mins. RNA contamination was removed by incubation with RNase and protein were digested with proteinase K. DNA was then extracted on a Promega Maxwell DX 16 for automated nucleic acid extraction using a Maxwell 16 LEV Blood DNA kit (Promega). Shotgun metagenomics library preparation and sequencing was performed by Genewiz using a NovaSeq platform (Illumina) obtaining sequence depth between 12.9 – 20 gb (43000000 – 69000000 reads per sample). Low quality reads and adaptors were removed using Trimmomatic v 0.69[91]. Kneaddata v0.73 (http://huttenhower.sph.harvard.edu/kneaddata) human and bacterial 16S sequence contaminants were removed and pair-end reads aligned. Bacterial taxonomic and functional identifications were determined using MetaPhlAn 2.0[92] and HUMAnN2[93] respectively. Bacterial taxa and functions that were not present in more than 40% of samples from one timepoint and with a total relative abundance less than 0.01% were removed for statistical analysis. Bacterial relative abundances were then log2 transformed. Two samples from one subject at baseline and W6 were not included in the metagenomic analysis as DNA failed to meet quality requirements.

### Multiplex assay

Plasma samples were isolated from blood collected at baseline, W6 and W12 (n = 20, 19 and 18, respectively) and inflammatory markers were analyzed by multiplex immunoassay using Bio-plex 200 system (Bio-rad, CA, USA) as per the manufacturer’s instructions. Circulating levels of cytokines were measured in duplicates by Bio-Plex Pro Human Cytokine 27-plex Assay (Catalog # M500KCAF0Y) including interleukin (IL)-1β, IL-1ra, IL-4, IL-7, IL-8 (chemokine CXCL8), macrophage inflammatory protein (MIP)-1α (CCL3), MIP-1β (CCL4), macrophage chemoattractant protein (MCP)-1 (CCL2), basic fibroblast growth factor (bFGF), interferon (IFN)-γ inducible protein (IP)-10 (CXCL10), IL-13, IL-9 and tumor necrosis factor (TNF)-α. C-peptide was measured using a Bio-Plex Pro^TM^ RBM Human Metabolic Panel 1 (Catalog # 171AMR1CK).

### RNA seq

Whole blood samples from participants were collected at baseline and W6 (n = 20 and 19, respectively) directly into Paxgene or Tempus tubes containing RNA stabilizers then stored at −80°C. RNA was purified according to the manufacturers’ instructions. Briefly, crude RNA was pelleted by centrifuging at 5000g for 15 minutes at 4°C. The pellet was washed, resuspended and treated with protease and DNase to remove contaminants. Finally, magnetic RNA-binding beads were added to capture RNA followed by additional washing and RNA elution. The purified RNA was checked for quality using Tapestation 4200 platform. Samples from two patients were not included in RNA-seq experiment due to the poor RNA integrity at either baseline or W6. Next-gene sequencing was performed by Genewiz using Illumina NovaSeq platform obtaining sequence depth of 50 million reads/sample. Transcriptome analysis was performed in samples from W0 and W6 for paired analysis.

### Statistics and reproducibility

Graphs were generated using R package ggplot2. Overall statistical significance was determined using geepack with pairwise differences identified by the package emmeans. *P < 0.05* was considered statistically significant. Total n refers to the number of biologically independent samples in each group. Box plots show mean and upper and lower quartile ranges and bar plots show mean and standard deviation. In more detail, changes in clinical parameters, safety, tolerability SCFAs, microbiota, immune phenotype and inflammatory measures over time were examined using generalized estimating equations (GEE) linear regression modelling unless otherwise stated. The GEE approach handles repeated measures and unbalanced designs whereby all observations with completed data at least one time point are included in analyses. All GEE models used robust standard errors estimation and repeated measurements over time were handled with unstructured correlation matrix. The linear regression assumptions of normality and homoscedasticity were checked prior to analyses to assess the suitability of a linear model. Analyses were performed in SPSS Version 25 software (IBM Corp. 1989, 2017).

For microbiota analysis, alpha diversity was calculated using the Inverse Simpson diversity index (Vegan, R package). Multivariate sparse partial least squares discriminant analyses accounting for repeated measures, quantified beta diversity between timepoints. Significant differences in beta diversity was determined using PERMANOVA (Adonis, R package). Differences in alpha diversity, taxa relative abundance and short chain fatty acids (acetate, butyrate and propionate) across timepoints were determined using generalized estimating equations (GEE), adjusting for multiple correction using the Benjamini-Hochberg approach (GeePack, R package). Estimated marginal means determined significant pairwise differences and includes a Tukey adjustment for multiple testing. Regressions across all timepoints between bacteria, SCFA’s and glycemic condition indices were performed using GEE generalized linearised modelling again adjusting for multiple correction using the Benjamini-Hochberg approach (GeePack, R package). Regression coefficient of zero indicates that the linear fit is worse than the null hypothesis of zero (or straight line). Correlations between glycemic condition and SCFA’s for each timepoint were performed using Pearson’s regression (R cran). Differences in percent live and mean signal intensity of immune populations across timepoints were determined using multivariate partial least squares discriminant analyses (PLS-DA), accounting for repeated measures. Significance was determined using PERMANOVA. The contribution of specific immune populations to the variation in immune markers between timepoints were determined by analyzing their contribution to the components in the PLS-DA and using univariate GEE analyses

For RNA-seq, statistical analysis of the of the quantified gene expression was performed using the edgeR v3.28.1[94] and RUVSeq v 1.0.0[95] Bioconductor packages within R statistical software. Differential expression analysis was performed comparing all samples at baseline with all samples at W6 taking account of the paired nature of the data and the tube used for sample collection (Paxgene or Tempus). The RUVSeq package was used to remove unwanted variation, employing the RUVg function and using a set of 1000 of the least variable genes as control genes. Gene set enrichment analysis was performed using the camera function of limma v. 3.42.2[96] to determine regulated pathways at W6.

## Data Availability

Metagenomic data is deposited in the European Nucleotide Archive (ENA) https://www.ebi.ac.uk/ena and RNA-seq data is uploaded in the Gene Expression Omnibus (GEO) database.

## Acknowledgements

We thank Dr Vivian Zhang, Kelsey Thompson, Curtis Huttenhower and the JDRF Microbiome Initiative Bioinformatics Center for assistance with metagenomics analysis. We would like to thank all the support staff at Sydney Cytometry and the Ramaciotti Facility for Human Systems Biology for their assistance with the mass cytometry studies. The bioinformatic analysis of gene expression data was supported in part by the Australian Government NCRIS program, administered by Bioplatforms Australia, and by the NSW State Government RAAP scheme.

## Author contributions

Author contributions: E.M. conceptualized, developed and lead the study, supervised S.B. and YA.Y., carried out investigation, wrote the manuscript and gave approval of the final version to be submitted. E.H-W. contributed to the study design, carried out investigation, supervised B.T and wrote the manuscript. E.M. and E.HW. take responsibility for the integrity of the data and the accuracy of data analysis. K.J.B co-designed the study, setup and conducted the clinical trial, analyzed data, supervised S.B., S.D., C.J.W. and wrote the manuscript. S.S. co-designed the study and carried out investigation. B.T. performed experiments, statistical analysis, and data visualisation, L.T.N, S.C. and M.W. contributed to analysis, interpretation and writing of the results. H.M.M., S.B. and YA.Y. and C.J.W performed experiments and contributed to analysis. S.B. and S.D. set-up and assisted in conducting the clinical trial and analyzing the dietary data. S.T. contributed to the design and interpretation and provided medical oversight. E.G., C.R.M., B.F. and L.H. contributed to the design, interpretation and writing.

## Availability of data

Metagenomic data is deposited in the European Nucleotide Archive (ENA) and RNA-seq data is uploaded in the GEO database. Data will be available upon publication.

## Declarations

The trial protocol was approved by the Sydney Local Health District Human Research Ethics Committee (HREC) (HREC/18/RPAH/241), and Monash University HREC. Clinical investigation of HAMSAB intervention was approved by Therapeutic Goods Administration (TGA) (CT-2020-CTN00691-1 v1). All patients provided written informed consent. The trial was registered with Australia New Zealand Clinical Trials Registry (ACTRN12618001391268).

## Consent for publication

Not Applicable.

## Supplementary information

**Fig. S1. sPLS-DA plot loadings indicating the contribution of each taxa.** Color corresponds to the timepoint in which the taxa is most abundant. Loadings are associated with Fig. 3B (taxa).

(A) Component 1 species distribution

(B) Component 2 species distribution

**Fig. S2. Gating Strategy for identification of immune cell phenotypes by CyTOF.**

**Fig. S3. Immune cell phenotype changes following HAMSAB treatment.**

(A) Log foldchange of frequency of live cells.

(B) Log foldchange of MSI from baseline. Asterisk represents GEE significance of changes in % live and MSI across timepoints: blue asterisk W0 vs W6 and red asterisk W0 vs W12. Mean and standard deviation are shown. #adjusted *P* < 0.1, *adjusted *P* = 0.1-0.05, **adjusted *P* = 0.05-0.001 ***adjusted *P* < 0.001.

**Fig. S4. Immune cell phenotype changes following HAMSAB treatment.**

Mass cytometry of PBMC was used to determine total frequency (within live cells) or MSI of:

(A) EM Tconv cells, Tconv TEMRA cells, CD8^+^ EM and CD8^+^ TEMRA cells.

(B) EM Tregs, TEMRA Tregs, naïve Tregs, CM Tregs.

(C) naïve and CM Tconv cells.

(D) Naïve and CM CD8^+^ T cells.

(E) EM and naïve Tconv expressing CTLA-4 and EM, naïve and TEMRA CD8^+^ T cells expressing CTLA4.

(F) Frequency of total pDCs, mDCs and MSI of CD86.

(G) Frequency of total non-classical, classical and intermediate monocytes. Colored dots represent each subject. Box plots show mean and upper and lower quartile ranges. Significance determined by GEE. Adjusted *P* values are (6W vs W0) or (12W vs W0). Gating strategy shown in Fig. S2.

**Fig. S5. Circulating pro-inflammatory and anti-inflammatory cytokines measured in subjects at baseline, W6 and W12 following HAMSAB supplementation.** Overall significance determined by GEE. Adjusted *P* >0.05 (not significant in all the groups). Coloured dots represent each subject. Box plots show mean and upper and lower quartile ranges.

**Fig. S6. Correlations between concentration of SCFAs, clinical parameters, relative abundance of bacterial communities and significant changes in immune cell subsets.**

(A) Pearson r values at each timepoint between plasma butyrate and stool acetate and butyrate with daily basal insulin.

(B) Pearson r values at each timepoint between plasma butyrate and stool acetate and butyrate with time below target continual glucose monitoring range (70 – 180 mg/dL). Grey shading represents 95% confidence intervals.

(C) GEE generalized linear modelling regressions accounting for repeated measures between Log2 relative abundances of *Bifidobacterium longum* and *adolescentis* and daily basal insulin.

(D) Regression modelling between frequency of total live immune cells with Log2 relative abundances of butyrate and acetate in plasma and stools using GEE generalized linearized modelling again adjusting for multiple correction using the Benjamini-Hochberg approach. Adjusted *P* values between the groups are shown on the graphs.

**Table S1. Nutritional and gastrointestinal changes across time.**

(A) Gastrointestinal and diabetes-specific quality of life assessments. Overall significance determined by GEE, adjusted *P* < 0.002. *P* < 0.003 (visit week 1 and week 4 vs baseline).

(B) Macronutrient intake across the study. Not significant.

**Table S2. Functional pathways encoded by the gut microbiota changed after taking dietary supplement.** Table of the 46 individual pathways that significantly differed across timepoints. The metaCyc superclasses assigned to each pathway are shown. Significance determined by GEE comparisons. Circles represent pairwise adjusted *P* values of either increases or decreases in abundance determined by estimated marginal means. Red indicates decrease, green indicates increase and black indicates no significant change. Small circle adjusted *P* value between 0.01 - 0.05, medium circle adjusted *P* value between 0.001 and 0.01, large circle adjusted *P* value is < 0.001.

**Table S3. Upregulated KEGG pathway gene sets identified using the camera function of limma following 6-weeks of HAMSAB supplementation.**

**Table S4. Results of differential gene expression testing with EdgeR and RUVseq comparing baseline and week 6.**

**Table S5. Mass cytometry panel used for immunophenotyping.**

## References

1. Thorburn AN, Macia L, Mackay CR: Diet, Metabolites, and “Western-Lifestyle” Inflammatory Diseases. Immunity 2014, 40(6):833–842.

2. Richards JL, Yap YA, McLeod KH, Mackay CR, Marino E: Dietary metabolites and the gut microbiota: an alternative approach to control inflammatory and autoimmune diseases. Clin Transl Immunology 2016, 5(5):e82.

3. de Goffau MC, Fuentes S, van den Bogert B, Honkanen H, de Vos WM, Welling GW, Hyoty H, Harmsen HJ: Aberrant gut microbiota composition at the onset of type 1 diabetes in young children. Diabetologia 2014, 57(8):1569–1577.

4. Zhao L, Zhang F, Ding X, Wu G, Lam YY, Wang X, Fu H, Xue X, Lu C, Ma J et al: Gut bacteria selectively promoted by dietary fibers alleviate type 2 diabetes. Science 2018, 359(6380):1151–1156.

5. Mariño E, Richards JL, McLeod KH, Stanley D, Yap YA, Knight J, McKenzie C, Kranich J, Oliveira AC, Rossello FJ et al: Gut microbial metabolites limit the frequency of autoimmune T cells and protect against type 1 diabetes. Nature immunology 2017, 18(5):552–562.

6. de Groot PF, Belzer C, Aydin O, Levin E, Levels JH, Aalvink S, Boot F, Holleman F, van Raalte DH, Scheithauer TP, et al: Distinct fecal and oral microbiota composition in human type 1 diabetes, an observational study. PLoS One 2017, 12(12):e0188475.

7. Vatanen T, Franzosa EA, Schwager R, Tripathi S, Arthur TD, Vehik K, Lernmark A, Hagopian WA, Rewers MJ, She JX et al: The human gut microbiome in early-onset type 1 diabetes from the TEDDY study. Nature 2018, 562(7728):589–594.

8. Huang J, Pearson JA, Peng J, Hu Y, Sha S, Xing Y, Huang G, Li X, Hu F, Xie Z et al: Gut microbial metabolites alter IgA immunity in type 1 diabetes. JCI Insight 2020.

9. Gavin PG, Mullaney JA, Loo D, Cao KL, Gottlieb PA, Hill MM, Zipris D, Hamilton-Williams EE: Intestinal Metaproteomics Reveals Host-Microbiota Interactions in Subjects at Risk for Type 1 Diabetes. Diabetes Care 2018, 41(10):2178–2186.

10. Yap YA, McLeod KH, McKenzie CI, Gavin PG, Davalos-Salas M, Richards JL, Moore RJ, Lockett TJ, Clarke JM, Eng VV et al: An acetate-yielding diet imprints an immune and anti-microbial programme against enteric infection. Clin Transl Immunology 2021, 10(1):e1233.

11. Roth-Schulze AJ, Penno MAS, Ngui KM, Oakey H, Bandala-Sanchez E, Smith AD, Allnutt TR, Thomson RL, Vuillermin PJ, Craig ME et al: Type 1 diabetes in pregnancy is associated with distinct changes in the composition and function of the gut microbiome. Microbiome 2021, 9(1):167.

12. Yap YA, Marino E: Dietary SCFAs Immunotherapy: Reshaping the Gut Microbiota in Diabetes. Adv Exp Med Biol 2020.

13. Mullaney JA, Stephens JE, Geeling BE, Hamilton-Williams EE: Early-life exposure to gut microbiota from disease-protected mice does not impact disease outcome in type 1 diabetes susceptible NOD mice. Immunol Cell Biol 2019, 97(1):97–103.

14. Wegh CAM, Geerlings SY, Knol J, Roeselers G, Belzer C: Postbiotics and Their Potential Applications in Early Life Nutrition and Beyond. Int J Mol Sci 2019, 20(19).

15. He Y, Fu L, Li Y, Wang W, Gong M, Zhang J, Dong X, Huang J, Wang Q, Mackay CR et al: Gut microbial metabolites facilitate anticancer therapy efficacy by modulating cytotoxic CD8(+) T cell immunity. Cell Metab 2021, 33(5):988–1000 e1007.

16. Fukuda S, Toh H, Hase K, Oshima K, Nakanishi Y, Yoshimura K, Tobe T, Clarke JM, Topping DL, Suzuki T et al: Bifidobacteria can protect from enteropathogenic infection through production of acetate. Nature 2011, 469(7331):543–547.

17. Stewart ML, Zimmer JP: Postprandial glucose and insulin response to a high-fiber muffin top containing resistant starch type 4 in healthy adults: a double-blind, randomized, controlled trial. Nutrition 2018, 53:59–63.

18. Wang Y, Chen J, Song YH, Zhao R, Xia L, Chen Y, Cui YP, Rao ZY, Zhou Y, Zhuang W et al: Effects of the resistant starch on glucose, insulin, insulin resistance, and lipid parameters in overweight or obese adults: a systematic review and meta-analysis. Nutr Diabetes 2019, 9(1):19.

19. Raghupathy P, Ramakrishna BS, Oommen SP, Ahmed MS, Priyaa G, Dziura J, Young GP, Binder HJ: Amylase-resistant starch as adjunct to oral rehydration therapy in children with diarrhea. J Pediatr Gastroenterol Nutr 2006, 42(4):362–368.

20. Aryana K, Greenway F, Dhurandhar N, Tulley R, Finley J, Keenan M, Martin R, Pelkman C, Olson D, Zheng J: A resistant-starch enriched yogurt: fermentability, sensory characteristics, and a pilot study in children. F1000Res 2015, 4:139.

21. Le Leu RK, Winter JM, Christophersen CT, Young GP, Humphreys KJ, Hu Y, Gratz SW, Miller RB, Topping DL, Bird AR et al: Butyrylated starch intake can prevent red meat-induced O6-methyl-2-deoxyguanosine adducts in human rectal tissue: a randomised clinical trial. Br J Nutr 2015:1–11.

22. Clarke JM, Topping DL, Christophersen CT, Bird AR, Lange K, Saunders I, Cobiac L: Butyrate esterified to starch is released in the human gastrointestinal tract. Am J Clin Nutr 2011, 94(5):1276–1283.

23. West NP, Christophersen CT, Pyne DB, Cripps AW, Conlon MA, Topping DL, Kang S, McSweeney CS, Fricker PA, Aguirre D et al: Butyrylated starch increases colonic butyrate concentration but has limited effects on immunity in healthy physically active individuals. Exercise immunology review 2013, 19:102–119.

24. Deehan EC, Yang C, Perez-Munoz ME, Nguyen NK, Cheng CC, Triador L, Zhang Z, Bakal JA, Walter J: Precision Microbiome Modulation with Discrete Dietary Fiber Structures Directs Short-Chain Fatty Acid Production. Cell Host Microbe 2020, 27(3):389–404 e386.

25. Braune A, Gutschow M, Blaut M: An NADH-Dependent Reductase from Eubacterium ramulus Catalyzes the Stereospecific Heteroring Cleavage of Flavanones and Flavanonols. Appl Environ Microbiol 2019, 85(19).

26. Chung WSF, Meijerink M, Zeuner B, Holck J, Louis P, Meyer AS, Wells JM, Flint HJ, Duncan SH: Prebiotic potential of pectin and pectic oligosaccharides to promote anti-inflammatory commensal bacteria in the human colon. FEMS Microbiol Ecol 2017, 93(11).

27. Swanson KS, de Vos WM, Martens EC, Gilbert JA, Menon RS, Soto-Vaca A, Hautvast J, Meyer PD, Borewicz K, Vaughan EE et al: Effect of fructans, prebiotics and fibres on the human gut microbiome assessed by 16S rRNA-based approaches: a review. Benef Microbes 2020, 11(2):101–129.

28. Wolfe AJ: The acetate switch. Microbiol Mol Biol Rev 2005, 69(1):12–50.

29. Bautista D, Vasquez C, Ayala-Ramirez P, Tellez-Sosa J, Godoy-Lozano E, Martinez-Barnetche J, Franco M, Angel J: Differential Expression of IgM and IgD Discriminates Two Subpopulations of Human Circulating IgM(+)IgD(+)CD27(+) B Cells That Differ Phenotypically, Functionally, and Genetically. Front Immunol 2020, 11:736.

30. Weller S, Braun MC, Tan BK, Rosenwald A, Cordier C, Conley ME, Plebani A, Kumararatne DS, Bonnet D, Tournilhac O et al: Human blood IgM “memory” B cells are circulating splenic marginal zone B cells harboring a prediversified immunoglobulin repertoire. Blood 2004, 104(12):3647–3654.

31. Mariño E, Batten M, Groom J, Walters S, Liuwantara D, Mackay F, Grey ST: Marginal-zone B-cells of nonobese diabetic mice expand with diabetes onset, invade the pancreatic lymph nodes, and present autoantigen to diabetogenic T-cells. Diabetes 2008, 57(2):395–404.

32. Tull TJ, Pitcher MJ, Guesdon W, Siu JHY, Lebrero-Fernandez C, Zhao Y, Petrov N, Heck S, Ellis R, Dhami P et al: Human marginal zone B cell development from early T2 progenitors. J Exp Med 2021, 218(4).

33. Cabrera SM, Engle S, Kaldunski M, Jia S, Geoffrey R, Simpson P, Szabo A, Speake C, Greenbaum CJ, Type 1 Diabetes TrialNet CISG et al: Innate immune activity as a predictor of persistent insulin secretion and association with responsiveness to CTLA4-Ig treatment in recent-onset type 1 diabetes. Diabetologia 2018, 61(11):2356–2370.

34. Joller N, Lozano E, Burkett PR, Patel B, Xiao S, Zhu C, Xia J, Tan TG, Sefik E, Yajnik V et al: Treg cells expressing the coinhibitory molecule TIGIT selectively inhibit proinflammatory Th1 and Th17 cell responses. Immunity 2014, 40(4):569–581.

35. Kolb H, Luckemeyer K, Heise T, Herder C, Schloot NC, Koenig W, Heinemann L, Martin S, Group DS: The systemic immune network in recent onset type 1 diabetes: central role of interleukin-1 receptor antagonist (DIATOR Trial). PLoS One 2013, 8(8):e72440.

36. Alnek K, Kisand K, Heilman K, Peet A, Varik K, Uibo R: Increased Blood Levels of Growth Factors, Proinflammatory Cytokines, and Th17 Cytokines in Patients with Newly Diagnosed Type 1 Diabetes. PLoS One 2015, 10(12):e0142976.

37. Furusawa Y, Obata Y, Fukuda S, Endo TA, Nakato G, Takahashi D, Nakanishi Y, Uetake C, Kato K, Kato T, et al: Commensal microbe-derived butyrate induces the differentiation of colonic regulatory T cells. Nature 2013, 504(7480):446–450.

38. Luu M, Pautz S, Kohl V, Singh R, Romero R, Lucas S, Hofmann J, Raifer H, Vachharajani N, Carrascosa LC et al: The short-chain fatty acid pentanoate suppresses autoimmunity by modulating the metabolic-epigenetic crosstalk in lymphocytes. Nat Commun 2019, 10(1):760.

39. Lochner M, Berod L, Sparwasser T: Fatty acid metabolism in the regulation of T cell function. Trends in immunology 2015, 36(2):81–91.

40. Yang Z, Matteson EL, Goronzy JJ, Weyand CM: T-cell metabolism in autoimmune disease. Arthritis Res Ther 2015, 17:29.

41. Maki KC, Pelkman CL, Finocchiaro ET, Kelley KM, Lawless AL, Schild AL, Rains TM: Resistant starch from high-amylose maize increases insulin sensitivity in overweight and obese men. The Journal of nutrition 2012, 142(4):717–723.

42. Giacco R, Parillo M, Rivellese AA, Lasorella G, Giacco A, D’Episcopo L, Riccardi G: Long-term dietary treatment with increased amounts of fiber-rich low-glycemic index natural foods improves blood glucose control and reduces the number of hypoglycemic events in type 1 diabetic patients. Diabetes Care 2000, 23(10):1461–1466.

43. Ho J, Nicolucci AC, Virtanen H, Schick A, Meddings J, Reimer RA, Huang C: Effect of Prebiotic on Microbiota, Intestinal Permeability, and Glycemic Control in Children With Type 1 Diabetes. The Journal of clinical endocrinology and metabolism 2019, 104(10):4427–4440.

44. de Groot PF, Nikolic T, Imangaliyev S, Bekkering S, Duinkerken G, Keij FM, Herrema H, Winkelmeijer M, Kroon J, Levin E et al: Oral butyrate does not affect innate immunity and islet autoimmunity in individuals with longstanding type 1 diabetes: a randomised controlled trial. Diabetologia 2020, 63(3):597–610.

45. Cleophas MCP, Ratter JM, Bekkering S, Quintin J, Schraa K, Stroes ES, Netea MG, Joosten LAB: Effects of oral butyrate supplementation on inflammatory potential of circulating peripheral blood mononuclear cells in healthy and obese males. Scientific reports 2019, 9(1):775.

46. den Besten G, van Eunen K, Groen AK, Venema K, Reijngoud DJ, Bakker BM: The role of short-chain fatty acids in the interplay between diet, gut microbiota, and host energy metabolism. J Lipid Res 2013, 54(9):2325–2340.

47. Donohoe DR, Garge N, Zhang X, Sun W, O’Connell TM, Bunger MK, Bultman SJ: The microbiome and butyrate regulate energy metabolism and autophagy in the mammalian colon. Cell Metabolism 2011, 13(5):517–526.

48. Donohoe DR, Collins LB, Wali A, Bigler R, Sun W, Bultman SJ: The Warburg effect dictates the mechanism of butyrate-mediated histone acetylation and cell proliferation. Molecular cell 2012, 48(4):612–626.

49. Dong W, Jia Y, Liu X, Zhang H, Li T, Huang W, Chen X, Wang F, Sun W, Wu H: Sodium butyrate activates NRF2 to ameliorate diabetic nephropathy possibly via inhibition of HDAC. J Endocrinol 2017, 232(1):71–83.

50. Bloemen JG, Venema K, van de Poll MC, Olde Damink SW, Buurman WA, Dejong CH: Short chain fatty acids exchange across the gut and liver in humans measured at surgery. Clinical nutrition (Edinburgh, Scotland) 2009, 28(6):657–661.

51. Moffett JR, Puthillathu N, Vengilote R, Jaworski DM, Namboodiri AM: Acetate Revisited: A Key Biomolecule at the Nexus of Metabolism, Epigenetics, and Oncogenesis – Part 2: Acetate and ACSS2 in Health and Disease. Frontiers in physiology 2020, 11(1451).

52. Richards RH, Vreman HJ, Zager P, Feldman C, Blaschke T, Weiner MW: Acetate Metabolism in Normal Human Subjects. American Journal of Kidney Diseases 1982, 2(1):47–57.

53. Chambers ES, Viardot A, Psichas A, Morrison DJ, Murphy KG, Zac-Varghese SE, MacDougall K, Preston T, Tedford C, Finlayson GS et al: Effects of targeted delivery of propionate to the human colon on appetite regulation, body weight maintenance and adiposity in overweight adults. Gut 2015, 64(11):1744–1754.

54. Belkaid Y, Harrison OJ: Homeostatic Immunity and the Microbiota. Immunity 2017, 46(4):562–576.

55. Yamamura R, Nakamura K, Kitada N, Aizawa T, Shimizu Y, Nakamura K, Ayabe T, Kimura T, Tamakoshi A: Associations of gut microbiota, dietary intake, and serum short-chain fatty acids with fecal short-chain fatty acids. Biosci Microbiota Food Health 2020, 39(1):11–17.

56. Juanola O, Ferrusquia-Acosta J, Garcia-Villalba R, Zapater P, Magaz M, Marin A, Olivas P, Baiges A, Bellot P, Turon F et al: Circulating levels of butyrate are inversely related to portal hypertension, endotoxemia, and systemic inflammation in patients with cirrhosis. FASEB journal : official publication of the Federation of American Societies for Experimental Biology 2019, 33(10):11595–11605.

57. Rosser EC, Oleinika K, Tonon S, Doyle R, Bosma A, Carter NA, Harris KA, Jones SA, Klein N, Mauri C: Regulatory B cells are induced by gut microbiota-driven interleukin-1beta and interleukin-6 production. Nat Med 2014, 20(11):1334–1339.

58. Daien CI, Tan J, Audo R, Mielle J, Quek LE, Krycer JR, Angelatos AS, Duares M, Pinget GV, Ni D, et al: Gut-derived acetate promotes B10 cells with anti-inflammatory effects. JCI Insight 2021.

59. So M, Elso CM, Tresoldi E, Pakusch M, Pathiraja V, Wentworth JM, Harrison LC, Krishnamurthy B, Thomas HE, Rodda C et al: Proinsulin C-peptide is an autoantigen in people with type 1 diabetes. Proc Natl Acad Sci U S A 2018, 115(42):10732–10737.

60. Musthaffa Y, Hamilton-Williams EE, Nel HJ, Bergot AS, Mehdi AM, Harris M, Thomas R: Proinsulin-specific T-cell responses correlate with estimated c-peptide and predict partial remission duration in type 1 diabetes. Clin Transl Immunology 2021, 10(7):e1315.

61. Diggins KE, Serti E, Muir V, Rosasco M, Lu T, Balmas E, Nepom G, Long SA, Linsley PS: Exhausted-like CD8+ T cell phenotypes linked to C-peptide preservation in alefacept-treated T1D subjects. JCI Insight 2021, 6(3).

62. Long SA, Thorpe J, DeBerg HA, Gersuk V, Eddy J, Harris KM, Ehlers M, Herold KC, Nepom GT, Linsley PS: Partial exhaustion of CD8 T cells and clinical response to teplizumab in new-onset type 1 diabetes. Sci Immunol 2016, 1(5).

63. Bolton HA, Zhu E, Terry AM, Guy TV, Koh WP, Tan SY, Power CA, Bertolino P, Lahl K, Sparwasser T et al: Selective Treg reconstitution during lymphopenia normalizes DC costimulation and prevents graft-versus-host disease. J Clin Invest 2015, 125(9):3627–3641.

64. Orban T, Beam CA, Xu P, Moore K, Jiang Q, Deng J, Muller S, Gottlieb P, Spain L, Peakman M et al: Reduction in CD4 central memory T-cell subset in costimulation modulator abatacept-treated patients with recent-onset type 1 diabetes is associated with slower C-peptide decline. Diabetes 2014, 63(10):3449–3457.

65. Narsale A, Davies JD: Memory T Cells in Type 1 Diabetes: the Devil is in the Detail. Current diabetes reports 2017, 17(8):61.

66. Devaraj S, Glaser N, Griffen S, Wang-Polagruto J, Miguelino E, Jialal I: Increased monocytic activity and biomarkers of inflammation in patients with type 1 diabetes. Diabetes 2006, 55(3):774–779.

67. Ren X, Mou W, Su C, Chen X, Zhang H, Cao B, Li X, Wu D, Ni X, Gui J et al: Increase in Peripheral Blood Intermediate Monocytes is Associated with the Development of Recent-Onset Type 1 Diabetes Mellitus in Children. Int J Biol Sci 2017, 13(2):209–218.

68. Irvine KM, Gallego P, An X, Best SE, Thomas G, Wells C, Harris M, Cotterill A, Thomas R: Peripheral blood monocyte gene expression profile clinically stratifies patients with recent-onset type 1 diabetes. Diabetes 2012, 61(5):1281–1290.

69. Van Sickle BJ, Simmons J, Hall R, Raines M, Ness K, Spagnoli A: Increased circulating IL-8 is associated with reduced IGF-1 and related to poor metabolic control in adolescents with type 1 diabetes mellitus. Cytokine 2009, 48(3):290–294.

70. Wang J, Ji H, Wang S, Liu H, Zhang W, Zhang D, Wang Y: Probiotic Lactobacillus plantarum Promotes Intestinal Barrier Function by Strengthening the Epithelium and Modulating Gut Microbiota. Front Microbiol 2018, 9:1953.

71. Chambers ES, Byrne CS, Morrison DJ, Murphy KG, Preston T, Tedford C, Garcia-Perez I, Fountana S, Serrano-Contreras JI, Holmes E et al: Dietary supplementation with inulin-propionate ester or inulin improves insulin sensitivity in adults with overweight and obesity with distinct effects on the gut microbiota, plasma metabolome and systemic inflammatory responses: a randomised cross-over trial. Gut 2019, 68(8):1430–1438.

72. Bhavsar I, Miller CS, Al-Sabbagh M: Macrophage inflammatory protein-1 Alpha (MIP-1 alpha)/CCL3: as a biomarker. General Methods in Biomarker Research and their Applications 2015:223.

73. Cameron MJ, Arreaza GA, Grattan M, Meagher C, Sharif S, Burdick MD, Strieter RM, Cook DN, Delovitch TL: Differential expression of CC chemokines and the CCR5 receptor in the pancreas is associated with progression to type I diabetes. The Journal of Immunology 2000, 165(2):1102–1110.

74. Nguyen NK, Deehan EC, Zhang Z, Jin M, Baskota N, Perez-Munoz ME, Cole J, Tuncil YE, Seethaler B, Wang T et al: Gut microbiota modulation with long-chain corn bran arabinoxylan in adults with overweight and obesity is linked to an individualized temporal increase in fecal propionate. Microbiome 2020, 8(1):118.

75. Oliphant K, Allen-Vercoe E: Macronutrient metabolism by the human gut microbiome: major fermentation by-products and their impact on host health. Microbiome 2019, 7(1):91.

76. Wang K, Liao M, Zhou N, Bao L, Ma K, Zheng Z, Wang Y, Liu C, Wang W, Wang J et al: Parabacteroides distasonis Alleviates Obesity and Metabolic Dysfunctions via Production of Succinate and Secondary Bile Acids. Cell Rep 2019, 26(1):222–235 e225.

77. Kverka M, Zakostelska Z, Klimesova K, Sokol D, Hudcovic T, Hrncir T, Rossmann P, Mrazek J, Kopecny J, Verdu EF et al: Oral administration of Parabacteroides distasonis antigens attenuates experimental murine colitis through modulation of immunity and microbiota composition. Clinical and experimental immunology 2011, 163(2):250–259.

78. Stewart CJ, Ajami NJ, O’Brien JL, Hutchinson DS, Smith DP, Wong MC, Ross MC, Lloyd RE, Doddapaneni H, Metcalf GA et al: Temporal development of the gut microbiome in early childhood from the TEDDY study. Nature 2018, 562(7728):583–588.

79. Zhou C, Zhao H, Xiao XY, Chen BD, Guo RJ, Wang Q, Chen H, Zhao LD, Zhang CC, Jiao YH, et al: Metagenomic profiling of the pro-inflammatory gut microbiota in ankylosing spondylitis. J Autoimmun 2020, 107:102360.

80. Belenguer A, Duncan SH, Calder AG, Holtrop G, Louis P, Lobley GE, Flint HJ: Two routes of metabolic cross-feeding between Bifidobacterium adolescentis and butyrate-producing anaerobes from the human gut. Appl Environ Microbiol 2006, 72(5):3593–3599.

81. Das P, Babaei P, Nielsen J: Metagenomic analysis of microbe-mediated vitamin metabolism in the human gut microbiome. BMC Genomics 2019, 20(1):208.

82. Hidalgo-Cantabrana C, Delgado S, Ruiz L, Ruas-Madiedo P, Sanchez B, Margolles A: Bifidobacteria and Their Health-Promoting Effects. Microbiol Spectr 2017, 5(3).

83. Sugahara H, Odamaki T, Fukuda S, Kato T, Xiao JZ, Abe F, Kikuchi J, Ohno H: Probiotic Bifidobacterium longum alters gut luminal metabolism through modification of the gut microbial community. Sci Rep 2015, 5:13548.

84. Yoshii K, Hosomi K, Sawane K, Kunisawa J: Metabolism of Dietary and Microbial Vitamin B Family in the Regulation of Host Immunity. Front Nutr 2019, 6:48.

85. Harbison JE, Roth-Schulze AJ, Giles LC, Tran CD, Ngui KM, Penno MA, Thomson RL, Wentworth JM, Colman PG, Craig ME et al: Gut microbiome dysbiosis and increased intestinal permeability in children with islet autoimmunity and type 1 diabetes: A prospective cohort study. Pediatric diabetes 2019, 20(5):574–583.

86. Secondulfo M, Iafusco D, Carratu R, deMagistris L, Sapone A, Generoso M, Mezzogiomo A, Sasso FC, Carteni M, De Rosa R et al: Ultrastructural mucosal alterations and increased intestinal permeability in non-celiac, type I diabetic patients. Dig Liver Dis 2004, 36(1):35–45.

87. Bosi E, Molteni L, Radaelli MG, Folini L, Fermo I, Bazzigaluppi E, Piemonti L, Pastore MR, Paroni R: Increased intestinal permeability precedes clinical onset of type 1 diabetes. Diabetologia 2006, 49(12):2824–2827.

88. Eypasch E, Williams JI, Wood-Dauphinee S, Ure BM, Schmulling C, Neugebauer E, Troidl H: Gastrointestinal Quality of Life Index: development, validation and application of a new instrument. Br J Surg 1995, 82(2):216–222.

89. Greenbaum CJ, Harrison LC, Immunology of Diabetes S: Guidelines for intervention trials in subjects with newly diagnosed type 1 diabetes. Diabetes 2003, 52(5):1059–1065.

90. Yu Z, Morrison M: Improved extraction of PCR-quality community DNA from digesta and fecal samples. Biotechniques 2004, 36(5):808–812.

91. Bolger AM, Lohse M, Usadel B: Trimmomatic: a flexible trimmer for Illumina sequence data. Bioinformatics 2014, 30(15):2114–2120.

92. Segata N, Waldron L, Ballarini A, Narasimhan V, Jousson O, Huttenhower C: Metagenomic microbial community profiling using unique clade-specific marker genes. Nat Methods 2012, 9(8):811–814.

93. Franzosa EA, McIver LJ, Rahnavard G, Thompson LR, Schirmer M, Weingart G, Lipson KS, Knight R, Caporaso JG, Segata N et al: Species-level functional profiling of metagenomes and metatranscriptomes. Nat Methods 2018, 15(11):962–968.

94. McCarthy DJ, Chen Y, Smyth GK: Differential expression analysis of multifactor RNA-Seq experiments with respect to biological variation. Nucleic Acids Res 2012, 40(10):4288–4297.

95. Risso D, Ngai J, Speed TP, Dudoit S: Normalization of RNA-seq data using factor analysis of control genes or samples. Nat Biotechnol 2014, 32(9):896–902.

96. Ritchie ME, Phipson B, Wu D, Hu Y, Law CW, Shi W, Smyth GK: limma powers differential expression analyses for RNA-sequencing and microarray studies. Nucleic Acids Res 2015, 43(7):e47.

