## Supplemental data for "Metabolite-based dietary supplementation in human type 1 diabetes is associated with microbiota and immune modulation"

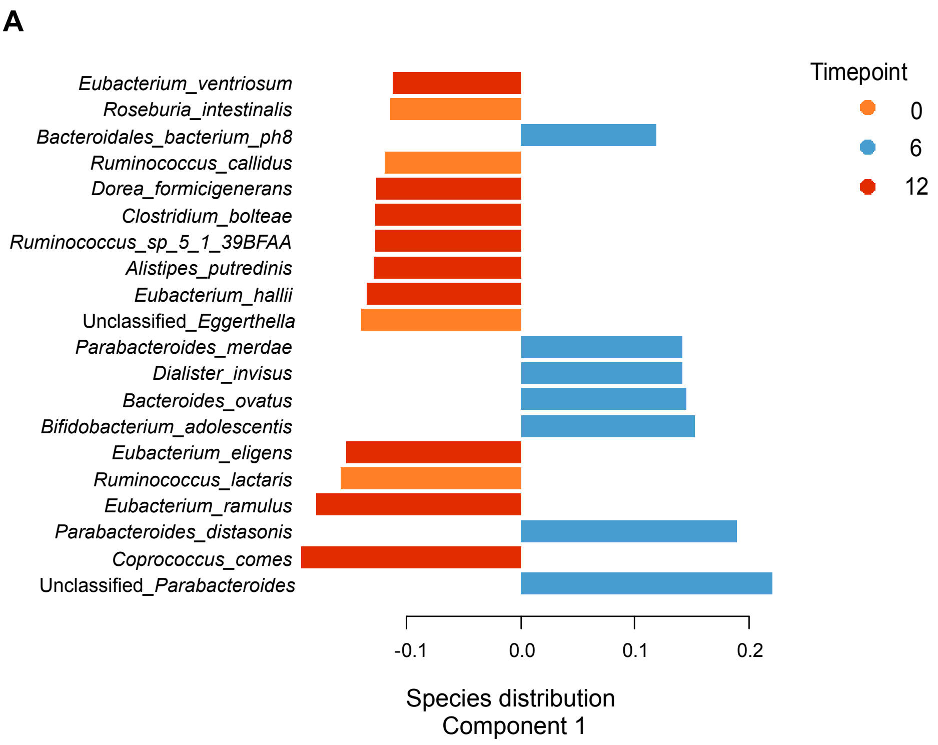

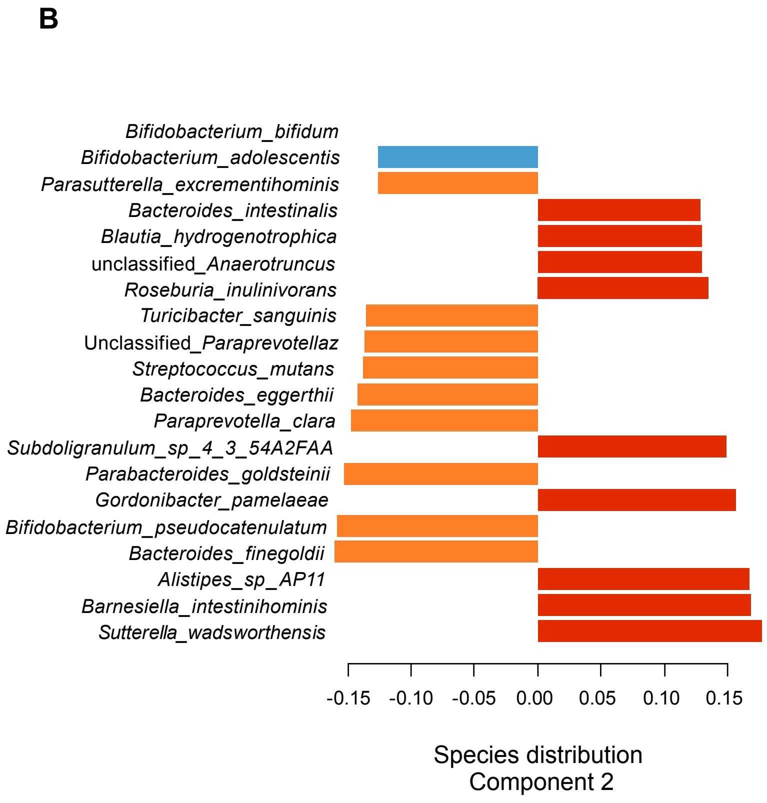

**Fig. S1. sPLS-DA plot loadings indicating the contribution of each taxa.** Color corresponds to the timepoint in which the taxa is most abundant. Loadings are associated with Fig. 3B (taxa).
(A) Component 1 species distribution
(B) Component 2 species distribution

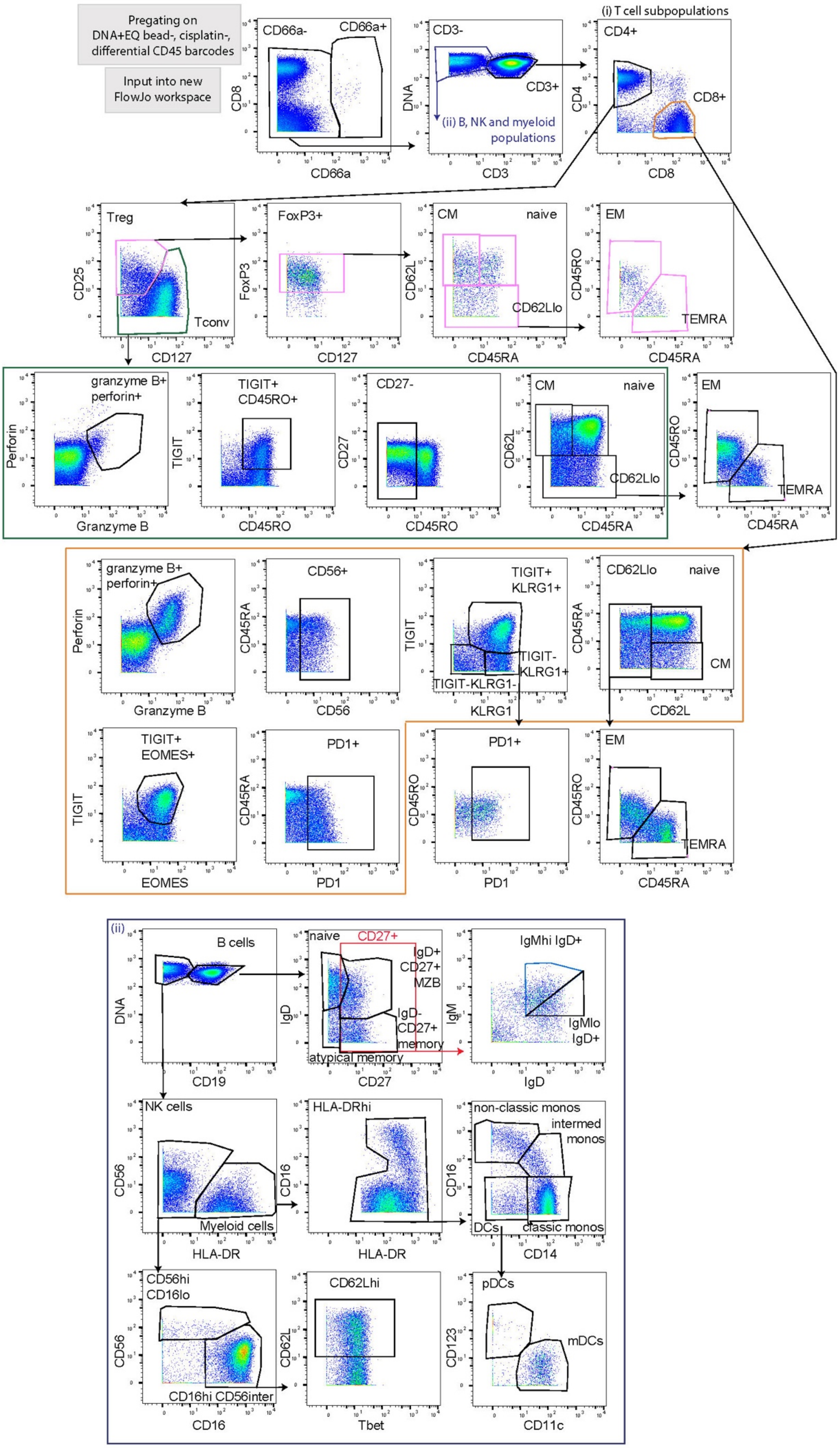

**Fig. S2. Gating Strategy for identification of immune cell phenotypes by CyTOF.**

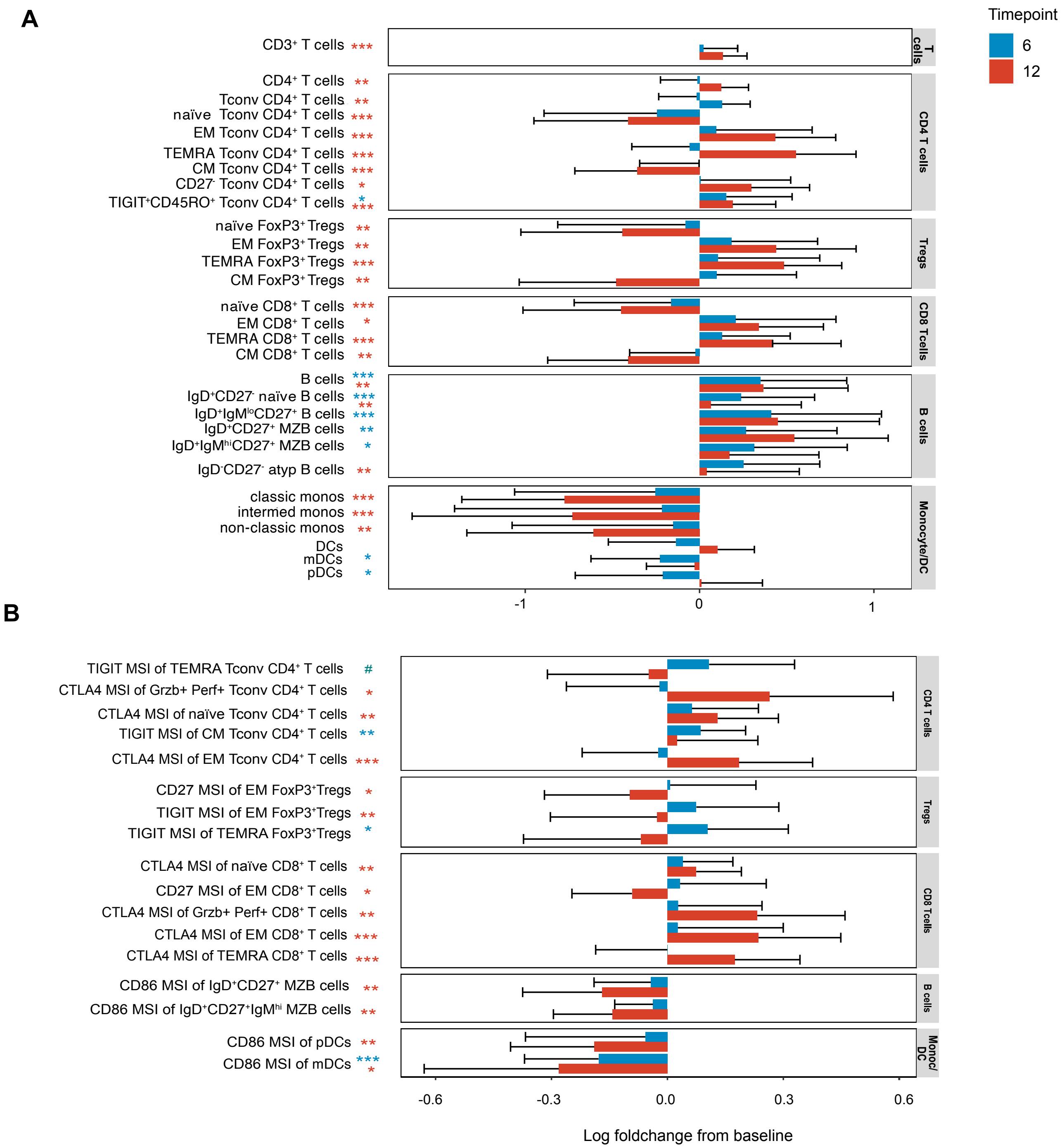

**Fig. S3. Immune cell phenotype changes following HAMSAB treatment.**
Log foldchange of frequency of live cells.
Log foldchange of MSI from baseline. Asterisk represents GEE significance of changes in % live and MSI across timepoints: blue asterisk W0 vs W6 and red asterisk W0 vs W12. Mean and standard deviation are shown. #adjusted $P < 0.1$, *adjusted $P = 0.1$- 0.05, **adjusted $P = 0.05$-0.001 ***adjusted $P < 0.001$.

*Bell, Saad & Tillett, et al., Fig. S4*

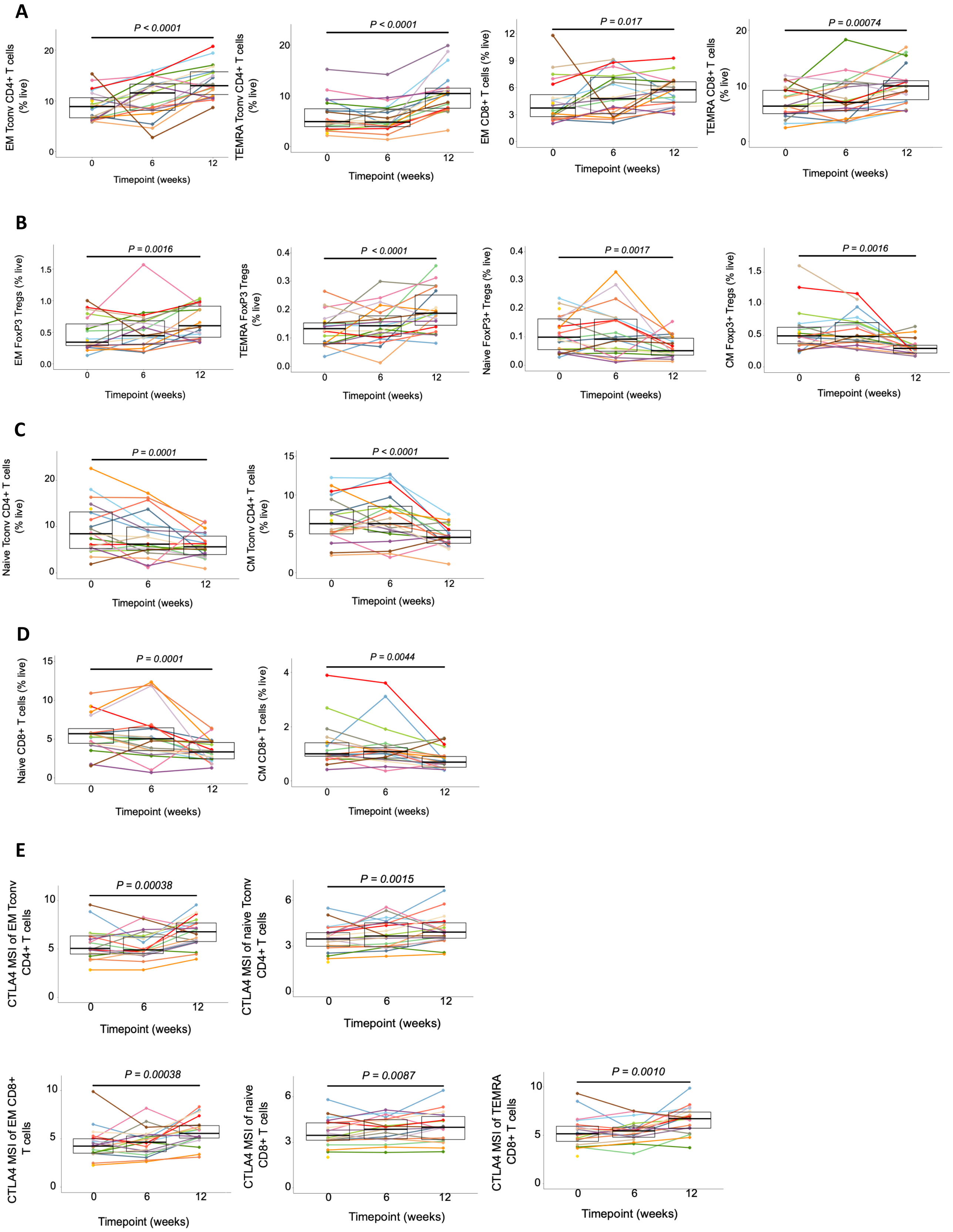

**F**

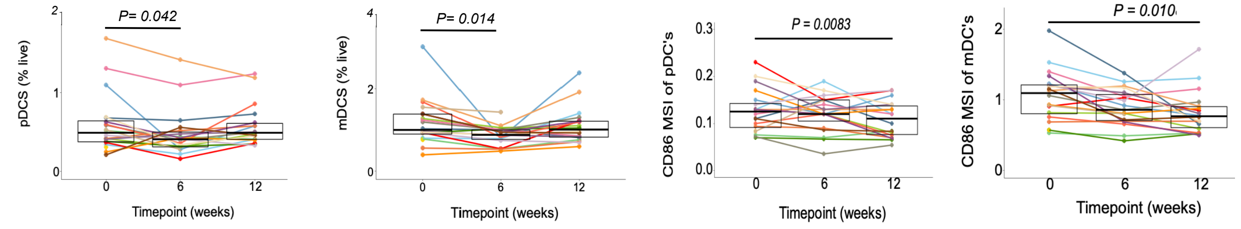

**G**

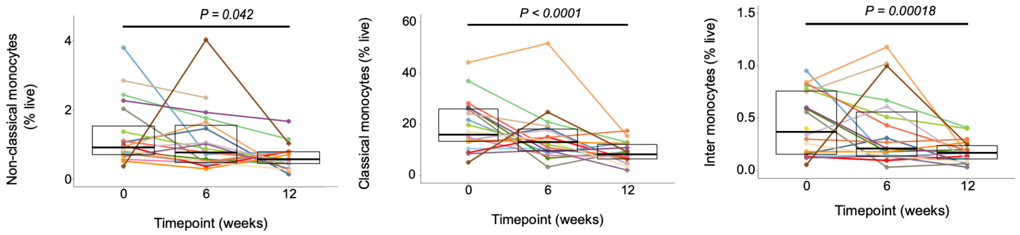

**Fig. S4. Immune cell phenotype changes following HAMSAB treatment.**
Mass cytometry of PBMC was used to determine total frequency (within live cells) or MSI of:
(A) EM Tconv cells, Tconv TEMRA cells, $CD8^+$ EM and $CD8^+$ TEMRA cells.
(B) EM Tregs, TEMRA Tregs, naïve Tregs, CM Tregs.
(C) naïve and CM Tconv cells.
(D) Naïve and CM $CD8^+$ T cells.
(E) EM and naïve Tconv expressing CTLA-4 and EM, naïve and TEMRA $CD8^+$ T cells expressing CTLA4.
(F) Frequency of total pDCs, mDCs and MSI of CD86.
(G) Frequency of total non-classical, classical and intermediate monocytes. Coloured dots represent each subject. Box plots show mean and upper and lower quartile ranges. Significance determined by GEE. Adjusted *P* values are (6W vs W0) or (12W vs W0). Gating strategy shown in Fig. S2.
.

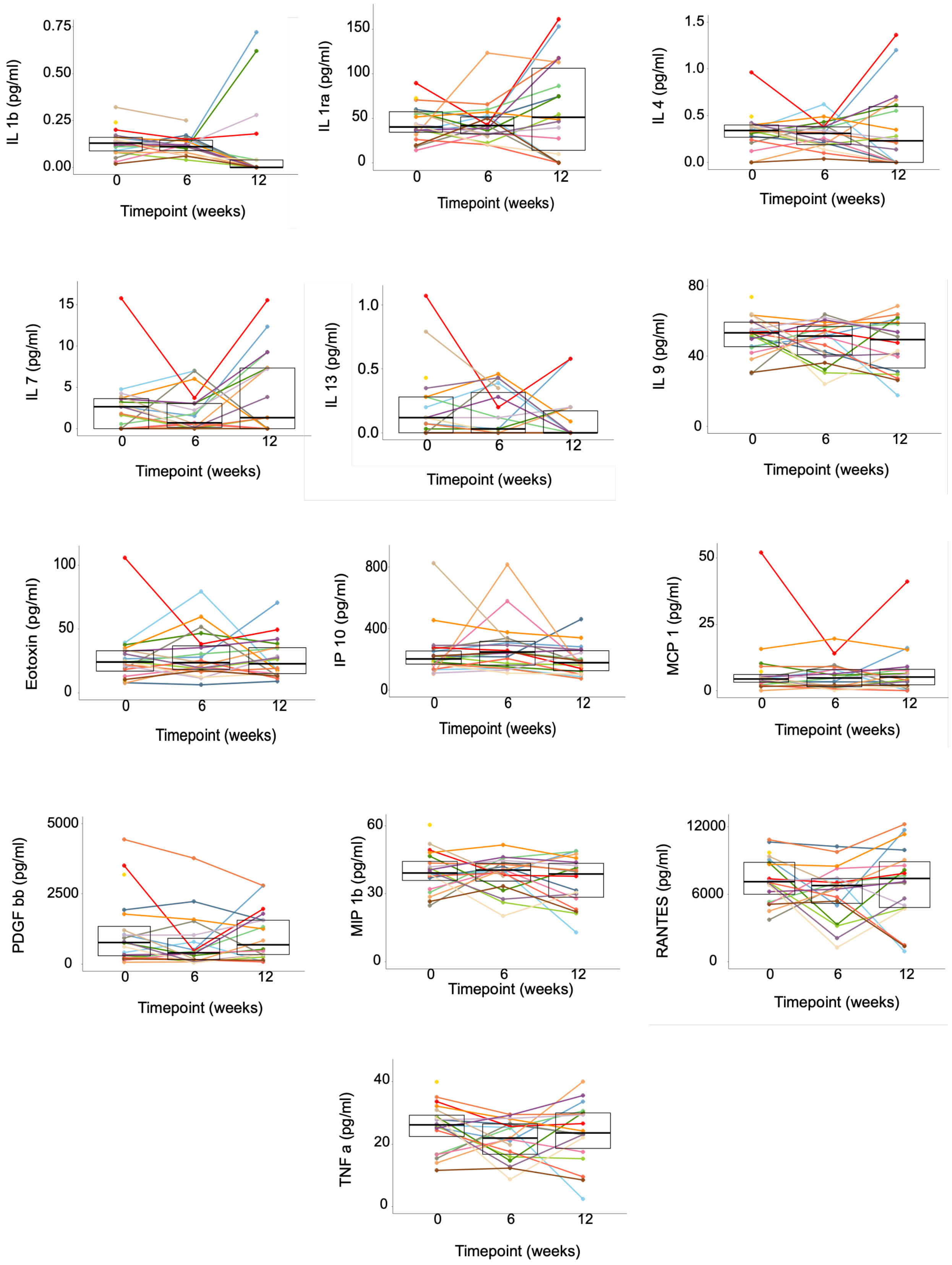

**Fig. S5. Circulating pro-inflammatory and anti-inflammatory cytokines measured in subjects at baseline, W6 and W12 following HAMSAB supplementation**. Overall significance determined by GEE. Adjusted *P* >0.05 (not significant in all the groups). Coloured dots represent each subject. Box plots show mean and upper and lower quartile ranges .

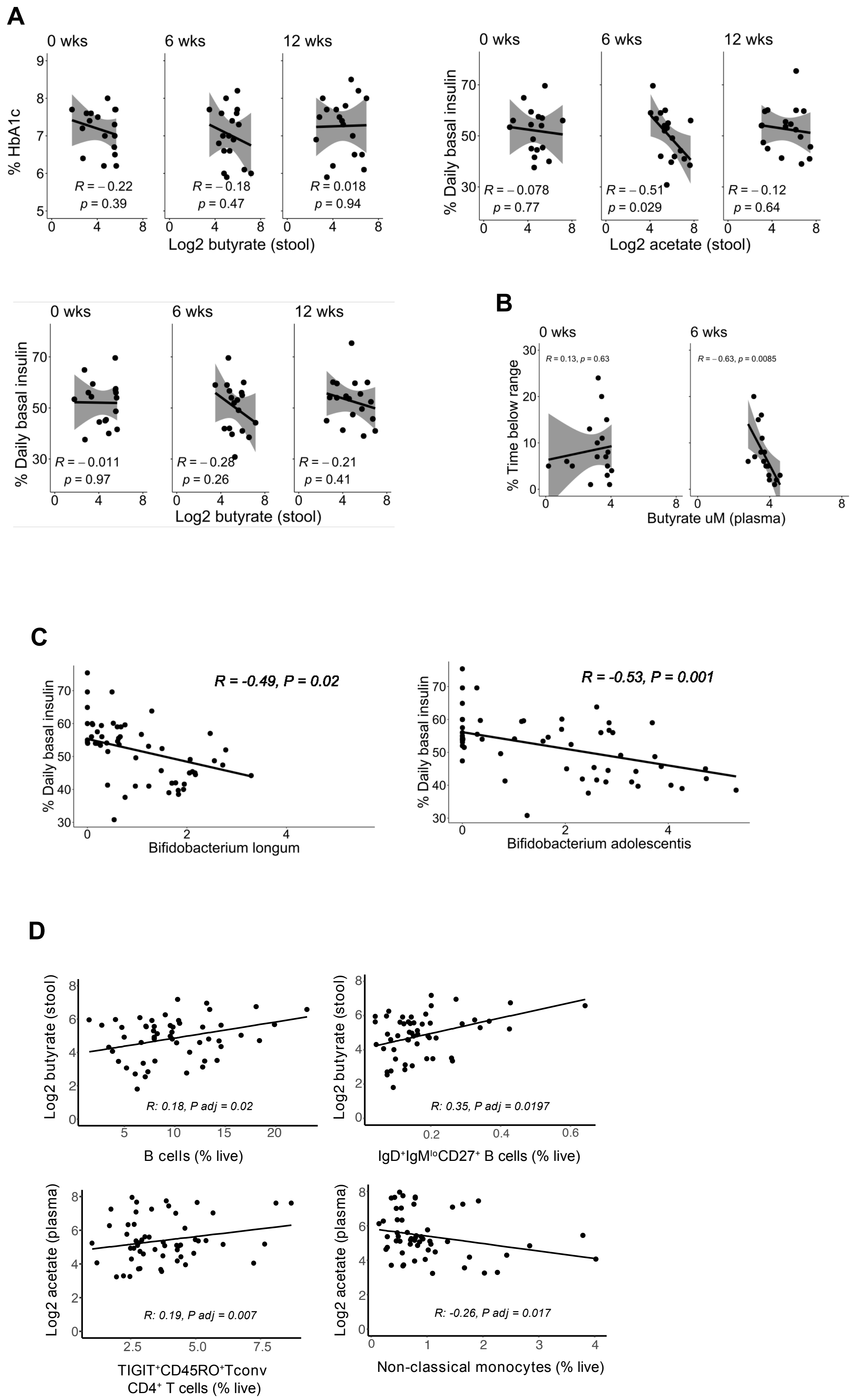
A
0 wks
6 wks
12 wks
% HbA1c
R = −0.22
p = 0.39
R = −0.18
p = 0.47
R = 0.018
p = 0.94
Log2 butyrate (stool)
% Daily basal insulin
R = −0.078
p = 0.77
R = −0.51
p = 0.029
R = −0.12
p = 0.64
Log2 acetate (stool)
R = −0.011
p = 0.97
R = −0.28
p = 0.26
R = −0.21
p = 0.41
Log2 butyrate (stool)
B
% Time below range
R = 0.13, p = 0.63
R = −0.63, p = 0.0085
Butyrate uM (plasma)
C
R = -0.49, P = 0.02
Bifidobacterium longum
R = -0.53, P = 0.001
Bifidobacterium adolescentis
D
Log2 butyrate (stool)
R: 0.18, P adj = 0.02
B cells (% live)
R: 0.35, P adj = 0.0197
IgD+IgMloCD27+ B cells (% live)
Log2 acetate (plasma)
R: 0.19, P adj = 0.007
TIGIT+CD45RO+Tconv
CD4+ T cells (% live)
R: -0.26, P adj = 0.017
Non-classical monocytes (% live)

**Fig. S6. Correlations between concentration of SCFAs, clinical parameters, relative abundance of bacterial communities and significant changes in immune cell subsets.**
(A) Pearson r values at each timepoint between plasma butyrate and stool acetate and butyrate with daily basal insulin.
(B) Pearson r values at each timepoint between plasma butyrate and stool acetate and butyrate with time below target continual glucose monitoring range (70 – 180 mg/dL). Grey shading represents 95% confidence intervals.
(C) GEE generalized linear modelling regressions accounting for repeated measures between Log2 relative abundances of *Bifidobacterium longum* and *adolescentis* and daily basal insulin. (D) Regression modelling between frequency of total live immune cells with Log2 relative abundances of butyrate and acetate in plasma and stools using GEE generalised linearised modelling again adjusting for multiple correction using the Benjamini-Hochberg approach. Adjusted *P* values between the groups are shown on the graphs .

### A. Gastrointestinal Symptoms and nutritional changes

| Questionnaires | Visit 1 (baseline) | Phone call (week 1) | Visit 2 (week 3) | Visit 3 (week 6) | Visit 4 (week 12) |
|---|---|---|---|---|---|
| **Gastrointestinal Quality of Life Index (GIQLI), (P = 0.002)** | 54 ± 6 (n=20) | 57 ± 4 (n=20), P = 0.003 | 55 ± 4 (n=20) | 54 ± 7 (n=20) | 57 ± 3 (n=19), P = 0.003 |
| **Diabetes-Specific Quality of Life** | 27 ± 5 (n=20) | 31 ± 3 (n=20) | 29 ± 4 (n=20) | 29 ± 5 (n=20) | 28 ± 6 (n=19) |

### B. Macronutrient intake did not differ between timepoints

| Nutrition Intake | Visit 1 (baseline) | Visit 3 (week 6) |
|---|---|---|
| **Energy (kJ [kCal])** | 8746 ± 2033 [2092 ± 486] (n=20) | 8686 ± 2469 [2078 ± 591] (n=18) |
| **Carbohydrate (% total energy intake)** | 37.3 ± 9.3 (n=20) | 37.0 ± 7.1 (n=18) |
| **Protein (% total energy intake)** | 15.9 ± 4.0 (n=20) | 16.7 ± 4.0 (n=18) |
| **Total Fat (% total energy intake)** | 41.0 ± 38.5 (n=20) | 38.5 ± 4.7 (n=18) |
| **Saturated Fat (% total energy intake)** | 14.5 ± 2.4 (n=20) | 14.3 ± 3.4 (n=18) |
| **Fibre* (g)** | 24.4 ± 7.0 (n=20) | 26.4 ± 10.3 (n=18) |
| **Glycemic Index (%)** | 54.2 ± 5.1 (n=20) | |
| **Glycemic Load (g)** | 108.6 ± 38.4 (n=20) | |

**Table S1. Nutritional and gastrointestinal changes across time.**
(A) Gastrointestinal and diabetes-specific quality of life assessments. Overall significance determined by GEE, adjusted *P* < 0.002. *P* < 0.003 (visit week 1 and week 4 vs baseline).
(B) Macronutrient intake across the study. Not significant.

| Pathway | Superclasses | Global FDR | p.value (0 - 6) | p.value (0 - 12) | p.value (6 - 12) |
|---|---|---|---|---|---|
| L.citrulline.biosynthesis | Amino.acid.biosynthesis | 0.0000 | green, large | black | red, large |
| Urea.cycle | Amino.acid.biosynthesis | 0.0000 | green, large | black | red, large |
| Calvin.Benson.Bassham.cycle | Carbohydrate.biosynthesis | 0.0000 | red, large | black | green, large |
| Pentose.phosphate.pathway (non.oxidative.branch) | Pentose.phosphate.pathways | 0.0000 | red, large | black | green, large |
| Glycolysis.II (fructose.6.phosphate) | Glycolysis | 0.0000 | green, large | black | red, large |
| Glycolysis I (glucose.6.phosphate) | Glycolysis | 0.0000 | green, large | black | red, large |
| Homolactic.fermentation | Fermentation | 0.0000 | green, large | black | black |
| Gluconeogenesis.III | Carbohydrate.biosynthesis | 0.0000 | green, large | black | red, large |
| Glycogen.degradation.II (eukaryotic) | Carbohydrate.degradation | 0.0001 | red, medium | black | green, large |
| Glycolysis.IV (plant.cytosol) | Glycolysis | 0.0039 | green, medium | black | red, medium |
| Acetyl.CoA.fermentation.to.butanoate.II | Fermentation | 0.0043 | red, medium | black | green, small |
| Glycolysis.VI (metazoan) | Glycolysis | 0.0094 | green, medium | black | red, small |
| Superpathway.of.hexitol.degradation (bacteria) | Secondary.metabolite.degradation | 0.0094 | green, medium | black | red, small |
| Chorismate.biosynthesis.from.3.dehydroquinate | Aromatic.compound.biosynthesis | 0.0118 | red, medium | black | green, small |
| GDP.mannose.biosynthesis | Carbohydrate.biosynthesis | 0.0118 | red, medium | black | black |
| Purine.nucleobases.degradation.I (anaerobic) | Nucleoside.Nucleotide.Degradation | 0.0118 | red, medium | black | black |
| Superpathway.of.pyrimidine.ribonucleosides.degradation | Nucleoside.Nucleotide.Degradation | 0.0118 | green, medium | black | red, small |
| Tetrapyrrole.biosynthesis.I (from.glutamate) | Tetrapyrrole.biosynthesis | 0.0118 | red, medium | black | black |
| L.rhamnose.degradation.I | Carbohydrate.degradation | 0.0159 | red, medium | black | green, medium |
| D.galactose.degradation.V (Leloir.pathway) | Carbohydrate.degradation | 0.0174 | red, medium | black | green, small |
| Dalactose.degradation.I (Leloir.pathway) | Carbohydrate.degradation | 0.0174 | red, medium | black | green, small |
| Guanosine.nucleotides.degradation.II | Nucleoside.Nucleotide.Degradation | 0.0246 | red, medium | black | black |
| Chorismate.biosynthesis.I | Aromatic.compound.biosynthesis | 0.0303 | red, medium | black | green, small |
| Creatinine.degradation.I | Amine.polyamine.biosynthesis | 0.0308 | red, medium | black | green, small |
| L.methionine.biosynthesis.III | Amino.acid.biosynthesis | 0.0325 | red, medium | black | green, small |
| Palmitoleate.biosynthesis.I (from..5Z..dodec.5.enoate) | Fatty.acid.lipid.biosynthesis | 0.0325 | green, medium | black | black |
| Superpathway.of.glycolysis.and.Entner.Doudoroff | Glycolysis | 0.0325 | green, small | black | black |
| Superpathway.of.arginine.and.polyamine.biosynthesis | Amine.polyamine.biosynthesis | 0.0332 | red, small | black | black |
| Gluconeogenesis.I | Carbohydrate.biosynthesis | 0.0337 | green, small | black | black |
| Superpathway.of..beta..D.glucuronide.and.D.glucuronate.degradation | Carboxylate.degradation | 0.0345 | red, small | black | black |
| Guanosine.nucleotides.degradation.III | Nucleoside.Nucleotide.Degradation | 0.0347 | red, small | black | black |
| Superpathway.of.L.serine.and.glycine.biosynthesis.I | Amino.acid.biosynthesis | 0.0357 | red, small | black | black |
| Superpathway.of.polyamine.biosynthesis.I | Amine.polyamine.biosynthesis | 0.0380 | red, small | black | black |
| Superpathway.of.pyridoxal.5 (phosphate.biosynthesis.and.salvage) | Cofactor.Carrier.Vitamin.biosynthesis | 0.0429 | green, small | black | red, small |
| Adenosine.nucleotides.degradation.II | Nucleoside.Nucleotide.Degradation | 0.0440 | red, small | black | green, small |
| 4.amino.2.methyl.5.phosphomethylpyrimidine.biosynthesis (yeast) | Cofactor.Carrier.Vitamin.biosynthesis | 0.0487 | green, small | black | black |
| 4.deoxy.L.threo.hex.4.enopyranuronate.degradation | Secondary.metabolite.degradation | 0.0487 | red, small | black | green, small |
| 5.aminoimidazole.ribonucleotide.biosynthesis.I | Nucleoside.Nucleotide.biosynthesis | 0.0487 | red, small | black | green, small |
| 5.aminoimidazole.ribonucleotide.biosynthesis.II | Nucleoside.Nucleotide.biosynthesis | 0.0487 | red, small | black | green, small |
| Biotin.biosynthesis.II | Cofactor.Carrier.Vitamin.biosynthesis | 0.0487 | green, small | black | red, small |
| Fatty.acid.elongation (saturated) | Fatty.acid.lipid.biosynthesis | 0.0487 | green, small | black | green, small |
| Flavin.biosynthesis.III | Cofactor.Carrier.Vitamin.biosynthesis | 0.0487 | green, small | black | red, small |
| Folate.transformations.II | Cofactor.Carrier.Vitamin.biosynthesis | 0.0487 | green, small | black | black |
| N10.formyl.tetrahydrofolate.biosynthesis | Cofactor.Carrier.Vitamin.biosynthesis | 0.0487 | green, small | black | red, small |
| Palmitate.biosynthesis.II (bacteria.and.plants) | Fatty.acid.lipid.biosynthesis | 0.0487 | black | black | green, small |
| Purine.ribonucleosides.degradation | Nucleoside.Nucleotide.Degradation | 0.0487 | black | black | green, small |

**Table S2. Functional pathways encoded by the gut microbiota changed after taking dietary supplement.** Table of the 46 individual pathways that significantly differed across timepoints. The metaCyc superclasses assigned to each pathway are shown. Significance determined by GEE comparisons. Circles represent pairwise adjusted *P* values of either increases or decreases in abundance determined by estimated marginal means. Red indicates decrease, green indicates increase and black indicates no significant change. Small circle adjusted *P* value between 0.01 - 0.05, medium circle adjusted *P* value between 0.001 and 0.01, large circle adjusted *P* value is < 0.001.

Extended Data Table 3: Upregulated KEGG pathway gene sets identified using thecamera function of limma following 6-weeks of HAMSAB supplementation

| | **NGenes** | **Direction** | **PValue** | **FDR** |
|---|---|---|---|---|
| KEGG_PATHOGENIC_ESCHERICHIA_COLI_INFECTION | 46 | Up | 1.03E-05 | **0.00032561** |
| KEGG_SPLICEOSOME | 123 | Up | 1.87E-05 | **0.00051181** |
| KEGG_ENDOCYTOSIS | 153 | Up | 2.09E-05 | **0.00054291** |
| KEGG_RIBOSOME | 85 | Up | 4.76E-05 | **0.0009854** |
| KEGG_PARKINSONS_DISEASE | 112 | Up | 0.00010368 | **0.0016533** |
| KEGG_TOLL_LIKE_RECEPTOR_SIGNALING_PATHWAY | 74 | Up | 0.00014285 | **0.0020007** |
| KEGG_ALZHEIMERS_DISEASE | 140 | Up | 0.00019282 | **0.00243343** |
| **KEGG_OXIDATIVE_PHOSPHORYLATION** | 114 | Up | 0.00022402 | **0.00268366** |
| KEGG_MELANOMA | 45 | Up | 0.00027273 | **0.0030361** |
| KEGG_LEISHMANIA_INFECTION | 62 | Up | 0.00029039 | **0.00317275** |
| KEGG_ANTIGEN_PROCESSING_AND_PRESENTATION | 59 | Up | 0.00033346 | **0.00342159** |
| KEGG_EPITHELIAL_CELL_SIGNALING_IN_HELICOBACTER_PYLORI_INFECTION | 55 | Up | 0.00070697 | **0.00570295** |
| KEGG_HUNTINGTONS_DISEASE | 152 | Up | 0.00076341 | **0.00602555** |
| KEGG_NOD_LIKE_RECEPTOR_SIGNALING_PATHWAY | 50 | Up | 0.00093818 | **0.00686541** |
| KEGG_RENAL_CELL_CARCINOMA | 60 | Up | 0.0010362 | **0.00742162** |
| KEGG_PROTEASOME | 41 | Up | 0.00109932 | **0.0076194** |
| KEGG_BASAL_TRANSCRIPTION_FACTORS | 27 | Up | 0.00126445 | **0.00845356** |
| KEGG_CHEMOKINE_SIGNALING_PATHWAY | 130 | Up | 0.00129201 | **0.00858914** |
| KEGG_NEUROTROPHIN_SIGNALING_PATHWAY | 105 | Up | 0.00144972 | **0.00932547** |
| KEGG_REGULATION_OF_ACTIN_CYTOSKELETON | 150 | Up | 0.00193589 | **0.01149356** |
| KEGG_CELL_ADHESION_MOLECULES_CAMS | 90 | Up | 0.00195375 | **0.01157105** |
| KEGG_FOCAL_ADHESION | 140 | Up | 0.00221472 | **0.01282635** |
| KEGG_CARDIAC_MUSCLE_CONTRACTION | 48 | Up | 0.00279329 | **0.01533061** |
| KEGG_NATURAL_KILLER_CELL_MEDIATED_CYTOTOXICITY | 94 | Up | 0.00291243 | **0.01574646** |

| | | | | |
|---|---|---|---|---|
| **KEGG_FATTY_ACID_METABOLISM** | 31 | Up | 0.0029675 | **0.01589851** |
| KEGG_BLADDER_CANCER | 35 | Up | 0.00302643 | **0.01615921** |
| KEGG_MAPK_SIGNALING_PATHWAY | 188 | Up | 0.00360128 | **0.01827748** |
| KEGG_GRAFT_VERSUS_HOST_DISEASE | 29 | Up | 0.00372 | **0.01873001** |
| KEGG_APOPTOSIS | 78 | Up | 0.00388889 | **0.01930131** |
| KEGG_RIG_I_LIKE_RECEPTOR_SIGNALING_PATHWAY | 50 | Up | 0.00419738 | **0.02048772** |
| KEGG_NUCLEOTIDE_EXCISION_REPAIR | 43 | Up | 0.00450588 | **0.02152606** |
| KEGG_P53_SIGNALING_PATHWAY | 58 | Up | 0.00469188 | **0.02214565** |
| KEGG_TYPE_I_DIABETES_MELLITUS | 31 | Up | 0.0049238 | **0.02287435** |
| KEGG_PANCREATIC_CANCER | 60 | Up | 0.00505389 | **0.02331806** |
| KEGG_GAP_JUNCTION | 62 | Up | 0.00526513 | **0.02405751** |
| KEGG_GLIOMA | 52 | Up | 0.00576047 | **0.02548211** |
| KEGG_UBIQUITIN_MEDIATED_PROTEOLYSIS | 128 | Up | 0.00673348 | **0.02852963** |
| KEGG_FC_GAMMA_R_MEDIATED_PHAGOCYTOSIS | 82 | Up | 0.00742183 | **0.03043531** |
| KEGG_PROSTATE_CANCER | 78 | Up | 0.0075133 | **0.03075697** |
| KEGG_LYSOSOME | 114 | Up | 0.00767149 | **0.03116128** |
| KEGG_LEUKOCYTE_TRANSENDOTHELIAL_MIGRATION | 77 | Up | 0.00785845 | **0.03158998** |
| KEGG_LONG_TERM_DEPRESSION | 45 | Up | 0.00832359 | **0.03282711** |
| KEGG_PATHWAYS_IN_CANCER | 232 | Up | 0.00923581 | **0.0354703** |
| KEGG_AUTOIMMUNE_THYROID_DISEASE | 29 | Up | 0.00938201 | **0.03582775** |
| KEGG_CYTOKINE_CYTOKINE_RECEPTOR_INTERACTION | 142 | Up | 0.00948482 | **0.03612752** |
| KEGG_O_GLYCAN_BIOSYNTHESIS | 19 | Up | 0.01145614 | **0.04146701** |
| KEGG_ALLOGRAFT_REJECTION | 28 | Up | 0.01161694 | **0.04174206** |
| KEGG_INSULIN_SIGNALING_PATHWAY | 109 | Up | 0.01168677 | **0.04190557** |
| KEGG_SYSTEMIC_LUPUS_ERYTHEMATOSUS | 72 | Up | 0.0122219 | **0.04305028** |
| KEGG_GLUTATHIONE_METABOLISM | 37 | Up | 0.01305531 | **0.04517673** |
| KEGG_ERBB_SIGNALING_PATHWAY | 68 | Up | 0.01450521 | **0.04835071** |
| KEGG_FC_EPSILON_RI_SIGNALING_PATHWAY | 60 | Up | 0.01461368 | **0.04854072** |

| | | | | |
|---|---|---|---|---|
| KEGG_PEROXISOME | 68 | Up | 0.01608822 | 0.05211834 |
| KEGG_HYPERTROPHIC_CARDIOMYOPATHY_HCM | 52 | Up | 0.01769088 | 0.05596578 |
| KEGG_PRION_DISEASES | 22 | Up | 0.01869297 | 0.05823816 |
| KEGG_HISTIDINE_METABOLISM | 17 | Up | 0.01985517 | 0.0607365 |
| KEGG_ASTHMA | 19 | Up | 0.02068978 | 0.06258573 |
| KEGG_PANTOTHENATE_AND_COA_BIOSYNTHESIS | 14 | Up | 0.02075427 | 0.06267445 |
| KEGG_AMYOTROPHIC_LATERAL_SCLEROSIS_ALS | 39 | Up | 0.03240687 | 0.08521562 |
| KEGG_VIRAL_MYOCARDITIS | 51 | Up | 0.03325894 | 0.08671254 |
| KEGG_ADIPOCYTOKINE_SIGNALING_PATHWAY | 52 | Up | 0.03549931 | 0.09071832 |
| KEGG_CHRONIC_MYELOID_LEUKEMIA | 65 | Up | 0.03662639 | 0.09294439 |
| KEGG_ARRHYTHMOGENIC_RIGHT_VENTRICULAR_CARDIOMYOPATHY_ARVC | 50 | Up | 0.03999511 | 0.09857803 |

**Extended Table 4: Results of differential gene expression testing with EdgeR and RUVseq comparing baseline and week 6.**

| | Symbol | Kegg | Entrez | EnsId | Biotype | logFC | logCPM | LR | PValue | FDR |
|---|---|---|---|---|---|---|---|---|---|---|
| ENSG00000163293 | NIPAL1 | K02927;K156 | 152519 | ENSG00000163293 | protein_coding | 0.89120734 | -0.2292553 | 21.431591 | 3.67E-06 | **0.02950508** |
| ENSG00000265830 | AL592188.7 | - | NA | NA | NA | -1.6461481 | 3.87234772 | 20.9804721 | 4.64E-06 | **0.02950508** |
| ENSG00000151789 | ZNF385D | - | 79750 | ENSG00000151789 | protein_coding | 0.97465004 | -0.1851283 | 20.7996992 | 5.10E-06 | **0.02950508** |
| ENSG00000256682 | TAS2R12 | - | NA | ENSG00000256682 | transcribed_processed_pseu | -0.8103476 | 0.13631038 | 20.1731733 | 7.07E-06 | **0.03069855** |
| ENSG00000244620 | AL122127.25 | - | NA | ENSG00000244620 | lncRNA | -0.8226105 | -0.2533075 | 17.9550127 | 2.26E-05 | 0.07852794 |
| ENSG00000234737 | KRT18P15 | K07604 | NA | ENSG00000234737 | processed_pseudogene | -0.7636077 | -0.0768158 | 16.3552774 | 5.25E-05 | 0.15191943 |
| ENSG00000200176 | RNU1-19P | - | NA | ENSG00000200176 | snRNA | -1.7081984 | -0.4483694 | 14.6018882 | 0.00013278 | 0.31017727 |
| ENSG00000262372 | RP11-669E14.6 | - | NA | ENSG00000262372 | lncRNA | -0.8576565 | 0.9211556 | 14.462905 | 0.00014295 | 0.31017727 |
| ENSG00000242330 | RN7SL683P | - | NA | ENSG00000242330 | misc_RNA | -0.8904398 | 0.58026012 | 13.7810519 | 0.0002054 | 0.32443031 |
| ENSG00000229531 | RP1-102G20.5 | - | NA | ENSG00000229531 | lncRNA | -0.9716506 | -0.6471222 | 13.6808892 | 0.00021665 | 0.32443031 |
| ENSG00000243151 | AL592188.1 | - | NA | NA | NA | -1.4369439 | 0.8413489 | 13.6594133 | 0.00021914 | 0.32443031 |
| ENSG00000213872 | AC092798.2 | K02912 | NA | ENSG00000213872 | processed_pseudogene | -0.5941114 | 1.27409491 | 13.2254338 | 0.00027618 | 0.32443031 |
| ENSG00000232150 | ST13P4 | K09560 | NA | ENSG00000232150 | processed_pseudogene | 0.79132674 | -0.0039007 | 13.2098057 | 0.00027849 | 0.32443031 |
| ENSG00000228939 | AKT3-IT1 | - | NA | ENSG00000228939 | lncRNA | -1.0941103 | 0.49959936 | 13.0622369 | 0.00030131 | 0.32443031 |
| ENSG00000230022 | FNTAP2 | K05955 | NA | ENSG00000230022 | processed_pseudogene | -0.6981356 | 0.2673223 | 12.9907356 | 0.00031304 | 0.32443031 |
| ENSG00000179639 | FCER1A | K08089 | 2205 | ENSG00000179639 | protein_coding | 0.69640723 | 5.57304406 | 12.9068765 | 0.00032738 | 0.32443031 |
| ENSG00000244926 | ALKBH3-AS1 | - | NA | ENSG00000244926 | lncRNA | -0.6325209 | 0.8576022 | 12.6807062 | 0.00036945 | 0.32443031 |
| ENSG00000227165 | WDR11-AS1 | - | NA | ENSG00000227165 | lncRNA | 0.64776202 | 1.82519829 | 12.6314678 | 0.00037931 | 0.32443031 |
| ENSG00000184261 | KCNK12 | K04921 | 56660 | ENSG00000184261 | protein_coding | -0.7086483 | 0.60782211 | 12.4976605 | 0.00040746 | 0.32443031 |
| ENSG00000213934 | HBG1 | K13824 | 3047 | ENSG00000213934 | protein_coding | 3.89714598 | 3.43276172 | 12.3404407 | 0.00044325 | 0.32443031 |
| ENSG00000259581 | TYRO3P | K05116 | NA | ENSG00000259581 | processed_pseudogene | -0.9149902 | 0.74110044 | 12.3397649 | 0.00044341 | 0.32443031 |
| ENSG00000199090 | MIR326 | - | NA | ENSG00000199090 | miRNA | -1.2076769 | -0.4754362 | 12.2335269 | 0.00046938 | 0.32443031 |
| ENSG00000264827 | AL592188.4 | - | NA | NA | NA | -1.2460294 | 0.0559869 | 12.1870994 | 0.00048121 | 0.32443031 |
| ENSG00000219712 | RP11-532F6.2 | - | NA | ENSG00000219712 | processed_pseudogene | -0.8229917 | 0.60213829 | 12.1421356 | 0.00049295 | 0.32443031 |
| ENSG00000236564 | YWHAQP5 | K01368 | NA | ENSG00000236564 | processed_pseudogene | -0.9190066 | 1.52792651 | 12.1135394 | 0.00050057 | 0.32443031 |
| ENSG00000232545 | KB-318B8.7 | - | NA | ENSG00000232545 | lncRNA | -0.8938741 | 0.80369888 | 12.0670249 | 0.00051322 | 0.32443031 |
| ENSG00000246214 | RP11-260E18.1 | - | NA | ENSG00000246214 | lncRNA | -0.8462127 | -0.3832809 | 12.0199871 | 0.00052633 | 0.32443031 |
| ENSG00000243423 | RP5-837J1.1 | K02922 | NA | ENSG00000243423 | processed_pseudogene | -0.7767684 | -0.0455007 | 12.0045989 | 0.00053069 | 0.32443031 |
| ENSG00000273245 | RP11-434P11.2 | - | NA | ENSG00000273245 | lncRNA | -0.9898235 | 0.68240356 | 11.8614905 | 0.00057307 | 0.32443031 |
| ENSG00000196565 | HBG2 | K13824 | 3048 | ENSG00000196565 | protein_coding | 2.18279037 | 3.63887166 | 11.8047345 | 0.0005908 | 0.32443031 |
| ENSG00000255139 | AP000442.1 | - | NA | ENSG00000255139 | lncRNA | -0.7669982 | -0.0717698 | 11.781517 | 0.00059822 | 0.32443031 |
| ENSG00000233668 | RP11-571F15.3 | K02995 | NA | ENSG00000233668 | processed_pseudogene | -0.7889548 | 0.26123087 | 11.7635972 | 0.000604 | 0.32443031 |
| ENSG00000231579 | RPL7P21 | - | NA | ENSG00000231579 | processed_pseudogene | -0.7188238 | 0.68129816 | 11.7247235 | 0.00061675 | 0.32443031 |
| ENSG00000169313 | P2RY12 | K04298 | 64805 | ENSG00000169313 | protein_coding | 0.66653542 | 3.47099239 | 11.5206187 | 0.00068828 | 0.35140987 |
| ENSG00000169224 | GCSAML | - | 148823 | ENSG00000169224 | protein_coding | 0.7473603 | 1.90500889 | 11.2952994 | 0.00077704 | 0.37209139 |
| ENSG00000114698 | PLSCR4 | - | 57088 | ENSG00000114698 | protein_coding | 0.8124095 | 0.3216348 | 11.2544785 | 0.00079431 | 0.37209139 |
| ENSG00000122872 | ARL4P | K07945 | NA | ENSG00000122872 | processed_pseudogene | -0.878271 | 0.01827066 | 11.2164842 | 0.00081074 | 0.37209139 |
| ENSG00000226394 | RP11-413E1.2 | K03948 | NA | ENSG00000226394 | processed_pseudogene | -0.7065362 | 1.06315787 | 11.188214 | 0.00082319 | 0.37209139 |
| ENSG00000226822 | RP11-356N1.2 | - | NA | ENSG00000226822 | lncRNA | -0.7504548 | 0.14859655 | 11.159626 | 0.00083597 | 0.37209139 |
| ENSG00000150681 | RGS18 | K16449 | 64407 | ENSG00000150681 | protein_coding | 0.57056757 | 7.82878097 | 11.110042 | 0.00085862 | 0.37261766 |
| ENSG00000267243 | AC005307.3 | - | NA | ENSG00000267243 | lncRNA | 0.78132256 | -0.2084932 | 11.0241719 | 0.00089931 | 0.37650406 |
| ENSG00000122043 | LINC00544 | - | NA | ENSG00000122043 | lncRNA | -0.6086489 | 0.24314712 | 10.9820301 | 0.00092 | 0.37650406 |
| ENSG00000222078 | RN7SKP110 | - | NA | ENSG00000222078 | misc_RNA | -0.8642628 | 0.58909339 | 10.9567377 | 0.00093264 | 0.37650406 |

| | | | | | | | | | | |
|---|---|---|---|---|---|---|---|---|---|---|
| ENSG00000262803 | RP11-160A9.3 | - | NA | ENSG00000262803 | processed_pseudogene | -0.6035765 | 0.59618593 | 10.7980811 | 0.00101605 | 0.39222015 |
| ENSG00000269999 | CTD-3185P2.2 | - | NA | NA | NA | -0.6058098 | 0.06204701 | 10.7967977 | 0.00101676 | 0.39222015 |
| ENSG00000242100 | RPL9P32 | K02940 | NA | ENSG00000242100 | processed_pseudogene | -1.1882256 | 0.3362082 | 10.6410903 | 0.00110602 | 0.39833901 |
| ENSG00000135525 | MAP7 | K10433 | 9053 | ENSG00000135525 | protein_coding | 0.5855958 | 2.34012891 | 10.6205574 | 0.00111837 | 0.39833901 |
| ENSG00000249693 | THEGL | - | 100506564 | ENSG00000249693 | protein_coding | 0.66788795 | -0.0567775 | 10.6137368 | 0.00112251 | 0.39833901 |
| ENSG00000270127 | RP11-526I2.5 | - | NA | ENSG00000270127 | lncRNA | -0.5692056 | 2.45076263 | 10.6106053 | 0.00112441 | 0.39833901 |
| ENSG00000140287 | HDC | K01590 | 3067 | ENSG00000140287 | protein_coding | 0.66616549 | 3.65920714 | 10.4632352 | 0.00121774 | 0.41460711 |
| ENSG00000236035 | RP11-90O23.1 | K14842 | NA | ENSG00000236035 | processed_pseudogene | -0.8446971 | 0.32008075 | 10.4626896 | 0.0012181 | 0.41460711 |
| ENSG00000168350 | DEGS2 | K04712 | 123099 | ENSG00000168350 | protein_coding | -0.5900463 | -0.228832 | 10.362431 | 0.00128606 | 0.42152871 |
| ENSG00000267529 | RP11-53B2.1 | - | NA | ENSG00000267529 | lncRNA | -0.5802076 | 1.44944206 | 10.3462963 | 0.00129735 | 0.42152871 |
| ENSG00000149534 | MS4A2 | K08090 | 2206 | ENSG00000149534 | protein_coding | 0.65017913 | 1.62677239 | 10.3265761 | 0.00131128 | 0.42152871 |
| ENSG00000090104 | RGS1 | K16449 | 5996 | ENSG00000090104 | protein_coding | 0.6924649 | 0.92880024 | 10.2270516 | 0.00138396 | 0.43542912 |
| ENSG00000257802 | MRS2P2 | K16075 | NA | ENSG00000257802 | processed_pseudogene | -0.5811773 | 1.39896949 | 10.1950498 | 0.00140818 | 0.43542912 |
| ENSG00000250030 | RP11-584P21.4 | K12581 | NA | ENSG00000250030 | processed_pseudogene | -0.7250826 | -0.2792747 | 10.0720188 | 0.00150538 | 0.43542912 |
| ENSG00000178033 | FAM26E | - | 254228 | ENSG00000178033 | protein_coding | 0.69042334 | 0.08536618 | 10.0586885 | 0.00151631 | 0.43542912 |
| ENSG00000235740 | RP11-436I24.1 | - | NA | ENSG00000235740 | lncRNA | -0.7618186 | 2.49456478 | 10.0552841 | 0.00151912 | 0.43542912 |
| ENSG00000180346 | TIGD2 | - | 166815 | ENSG00000180346 | protein_coding | 0.38947263 | 2.11125121 | 9.93964099 | 0.00161757 | 0.43542912 |
| ENSG00000121769 | FABP3 | K08752 | 2170 | ENSG00000121769 | protein_coding | -0.6654635 | 0.64344034 | 9.80970354 | 0.00173593 | 0.43542912 |
| ENSG00000272412 | Metazoa_SRP | - | NA | ENSG00000272412 | misc_RNA | -0.9478426 | -0.7710013 | 9.78658443 | 0.0017579 | 0.43542912 |
| ENSG00000133477 | FAM83F | K12581;K029 | 113828 | ENSG00000133477 | protein_coding | 0.74554827 | 0.32466309 | 9.78568989 | 0.00175875 | 0.43542912 |
| ENSG00000146122 | DAAM2 | K04512 | 23500 | ENSG00000146122 | protein_coding | 0.75304352 | 1.1087293 | 9.75941283 | 0.00178407 | 0.43542912 |
| ENSG00000242609 | RP11-398A8.1 | - | NA | ENSG00000242609 | processed_pseudogene | -0.943753 | -0.2999307 | 9.70985606 | 0.00183282 | 0.43542912 |
| ENSG00000244556 | ODCP | K01581 | NA | ENSG00000244556 | processed_pseudogene | -0.6129997 | 1.39159499 | 9.67699441 | 0.0018659 | 0.43542912 |
| ENSG00000270772 | RP11-332H21.2 | K00161 | NA | ENSG00000270772 | processed_pseudogene | -0.7537208 | 0.75104364 | 9.66135551 | 0.00188185 | 0.43542912 |
| ENSG00000244071 | RPL9P33 | - | NA | ENSG00000244071 | processed_pseudogene | -0.9680644 | 0.55139976 | 9.61127797 | 0.00193386 | 0.43542912 |
| ENSG00000148498 | PARD3 | K04237 | 56288 | ENSG00000148498 | protein_coding | 0.62730521 | 2.39204056 | 9.58654404 | 0.00196009 | 0.43542912 |
| ENSG00000214283 | RP11-85F14.1 | - | NA | ENSG00000214283 | processed_pseudogene | -0.7851905 | 0.50127068 | 9.55691682 | 0.00199197 | 0.43542912 |
| ENSG00000225213 | RP11-197M22.2 | - | NA | ENSG00000225213 | lncRNA | -0.9126482 | 0.00328871 | 9.50683701 | 0.00204708 | 0.43542912 |
| ENSG00000224072 | RP11-75A9.3 | - | NA | ENSG00000224072 | processed_pseudogene | -0.5018623 | 1.21348529 | 9.47028445 | 0.00208827 | 0.43542912 |
| ENSG00000200156 | RNU5B-1 | - | NA | ENSG00000200156 | snRNA | -1.9344189 | 3.05544087 | 9.46234796 | 0.00209732 | 0.43542912 |
| ENSG00000160193 | WDR4 | K15443 | 10785 | ENSG00000160193 | protein_coding | -0.6018569 | 2.55334198 | 9.44819244 | 0.00211357 | 0.43542912 |
| ENSG00000238961 | SNORA47 | - | NA | ENSG00000238961 | snoRNA | 0.54676923 | 1.63270152 | 9.41221088 | 0.00215545 | 0.43542912 |
| ENSG00000143429 | AC027612.6 | K14957 | NA | ENSG00000143429 | transcribed_unprocessed_ps | 0.63456364 | -0.0235772 | 9.40603674 | 0.00216272 | 0.43542912 |
| ENSG00000266222 | RP11-433M22.1 | - | NA | ENSG00000266222 | lncRNA | -1.3786895 | -0.5765254 | 9.39986868 | 0.00217001 | 0.43542912 |
| ENSG00000147650 | LRP12 | K12581;K092 | 29967 | ENSG00000147650 | protein_coding | 0.46231259 | 2.05756168 | 9.39796876 | 0.00217226 | 0.43542912 |
| ENSG00000264878 | Z85986.1 | - | NA | NA | NA | -0.8204803 | 0.64166997 | 9.39637173 | 0.00217415 | 0.43542912 |
| ENSG00000105971 | CAV2 | K12958 | 858 | ENSG00000105971 | protein_coding | 0.89624386 | 0.56507987 | 9.39170381 | 0.0021797 | 0.43542912 |
| ENSG00000169902 | TPST1 | K01021 | 8460 | ENSG00000169902 | protein_coding | 0.63024029 | 3.76623831 | 9.36097355 | 0.00221654 | 0.43542912 |
| ENSG00000259165 | DDX18P1 | K13179 | NA | ENSG00000259165 | processed_pseudogene | -0.4794796 | 2.10319687 | 9.26691834 | 0.0023333 | 0.43542912 |
| ENSG00000213216 | RP11-355O1.7 | K02975 | NA | ENSG00000213216 | processed_pseudogene | -0.5374137 | -0.3541348 | 9.23924616 | 0.00236882 | 0.43542912 |
| ENSG00000134317 | GRHL1 | K09275 | 29841 | ENSG00000134317 | protein_coding | 0.7113523 | 1.50511366 | 9.22652942 | 0.00238533 | 0.43542912 |
| ENSG00000180822 | PSMG4 | K11878;K114 | 389362 | ENSG00000180822 | protein_coding | -0.5626576 | 4.03413138 | 9.20162623 | 0.002418 | 0.43542912 |
| ENSG00000261096 | RP11-690I21.2 | - | NA | ENSG00000261096 | lncRNA | -0.9275259 | 0.29512228 | 9.19237684 | 0.00243025 | 0.43542912 |
| ENSG00000259781 | RP11-673C5.1 | K10802 | NA | ENSG00000259781 | processed_pseudogene | 0.55768482 | 2.01003051 | 9.1814456 | 0.00244481 | 0.43542912 |
| ENSG00000258839 | MC1R | K04199 | 4157 | ENSG00000258839 | protein_coding | -0.4984494 | 1.88646697 | 9.14375169 | 0.00249569 | 0.43542912 |
| ENSG00000241230 | RN7SL801P | - | NA | ENSG00000241230 | misc_RNA | -0.9994051 | -0.8719354 | 9.13207818 | 0.00251166 | 0.43542912 |

| | | | | | | | | | | | |
|---|---|---|---|---|---|---|---|---|---|---|---|
| ENSG00000225084 | AL450226.2 | - | NA | | ENSG00000225084 | lncRNA | -1.0944883 | -0.6367515 | 9.12872184 | 0.00251627 | 0.43542912 |
| ENSG00000233387 | RP11-342D11.3 | - | NA | | ENSG00000233387 | lncRNA | -0.5962417 | 1.01611416 | 9.11643451 | 0.00253323 | 0.43542912 |
| ENSG00000218313 | RP11-393I2.2 | K03004 | NA | | ENSG00000218313 | processed_pseudogene | -0.7196903 | 2.27709968 | 9.10218276 | 0.00255305 | 0.43542912 |
| ENSG00000206172 | HBA1 | K13822 | | 3039 | ENSG00000206172 | protein_coding | 2.57425627 | 6.79071081 | 9.1016418 | 0.0025538 | 0.43542912 |
| ENSG00000252949 | AL136303.1 | - | NA | | NA | NA | -0.7734006 | -0.0439744 | 9.09317747 | 0.00256565 | 0.43542912 |
| ENSG00000186049 | KRT73 | K07605 | | 319101 | ENSG00000186049 | protein_coding | -0.6310341 | 1.3030057 | 9.05516224 | 0.00261954 | 0.43542912 |
| ENSG00000137331 | IER3 | - | | 8870 | ENSG00000137331 | protein_coding | 0.57824846 | 2.48659946 | 9.03098965 | 0.00265441 | 0.43542912 |
| ENSG00000270711 | RP11-46A10.8 | - | NA | | ENSG00000270711 | processed_pseudogene | -0.5642782 | -0.096552 | 9.02601523 | 0.00266164 | 0.43542912 |
| ENSG00000259073 | FOXN3-AS2 | - | NA | | ENSG00000259073 | lncRNA | -0.9905673 | 1.93736594 | 9.01068733 | 0.00268405 | 0.43542912 |
| ENSG00000198829 | SUCNR1 | K10042 | | 56670 | ENSG00000198829 | protein_coding | 0.69733164 | 1.00221041 | 8.98718661 | 0.00271879 | 0.43542912 |
| ENSG00000233337 | UBE2FP3 | K10687 | NA | | ENSG00000233337 | processed_pseudogene | -0.555005 | 0.63784693 | 8.97256292 | 0.00274064 | 0.43542912 |
| ENSG00000175105 | ZNF654 | - | | 55279 | ENSG00000175105 | protein_coding | 0.41828049 | 5.20917483 | 8.96892697 | 0.0027461 | 0.43542912 |
| ENSG00000260536 | RP5-1085F17.4 | - | NA | | ENSG00000260536 | lncRNA | -0.6386072 | -0.0867436 | 8.95953932 | 0.00276024 | 0.43542912 |
| ENSG00000225338 | RP11-384C4.3 | K02893 | NA | | ENSG00000225338 | processed_pseudogene | -0.7379865 | 0.9478958 | 8.94833201 | 0.00277723 | 0.43542912 |
| ENSG00000225135 | RP11-361F15.2 | - | NA | | NA | NA | 0.57893038 | 3.79132538 | 8.92457141 | 0.00281359 | 0.43542912 |
| ENSG00000248477 | RP11-848G14.2 | K12807 | NA | | ENSG00000248477 | transcribed_unprocessed_ps | -0.601899 | -0.0471494 | 8.9220552 | 0.00281747 | 0.43542912 |
| ENSG00000041353 | RAB27B | K07886 | | 5874 | ENSG00000041353 | protein_coding | 0.53812147 | 6.06263225 | 8.92148931 | 0.00281835 | 0.43542912 |
| ENSG00000251194 | RP1-68D18.2 | - | NA | | ENSG00000251194 | lncRNA | -0.4370391 | 2.39129262 | 8.91889817 | 0.00282235 | 0.43542912 |
| ENSG00000254556 | AF131215.4 | - | NA | | ENSG00000254556 | lncRNA | -0.9212989 | 1.6966818 | 8.91199385 | 0.00283304 | 0.43542912 |
| ENSG00000236086 | HMGN2P28 | K11300 | NA | | ENSG00000236086 | processed_pseudogene | -0.5542456 | -0.1943794 | 8.89532003 | 0.00285902 | 0.43542912 |
| ENSG00000116918 | TSNAX | K17985 | | 7257 | ENSG00000116918 | protein_coding | 0.39627946 | 5.54421015 | 8.89500285 | 0.00285952 | 0.43542912 |
| ENSG00000154188 | ANGPT1 | K05465 | | 284 | ENSG00000154188 | protein_coding | 0.6714805 | 3.10272011 | 8.85886152 | 0.00291669 | 0.43542912 |
| ENSG00000259274 | CTD-2501E16.2 | - | NA | | ENSG00000259274 | lncRNA | -0.5521034 | 0.60660058 | 8.81787581 | 0.00298294 | 0.43542912 |
| ENSG00000151023 | ENKUR | - | | 219670 | ENSG00000151023 | protein_coding | 0.59257685 | 2.70603105 | 8.80094614 | 0.00301074 | 0.43542912 |
| ENSG00000237672 | KRR1P1 | K06961 | NA | | ENSG00000237672 | processed_pseudogene | -0.571237 | 1.24881057 | 8.79756975 | 0.00301632 | 0.43542912 |
| ENSG00000268362 | CTD-2017D11.1 | - | NA | | ENSG00000268362 | lncRNA | -0.6276071 | 3.35680106 | 8.76225313 | 0.00307529 | 0.43542912 |
| ENSG00000272486 | RP11-532M24.1 | - | NA | | NA | NA | -0.5946875 | 0.33231092 | 8.75490148 | 0.00308771 | 0.43542912 |
| ENSG00000119862 | LGALSL | - | | 29094 | ENSG00000119862 | protein_coding | 0.46801104 | 4.56850063 | 8.73132177 | 0.0031279 | 0.43542912 |
| ENSG00000181227 | RP4-682C21.2 | K00658 | NA | | ENSG00000181227 | processed_pseudogene | -0.9470581 | 0.49666119 | 8.72546921 | 0.00313795 | 0.43542912 |
| ENSG00000231697 | NANOGP5 | K10164 | NA | | ENSG00000231697 | transcribed_processed_pseu | -0.6068257 | 0.17112729 | 8.71599316 | 0.00315431 | 0.43542912 |
| ENSG00000255320 | RP11-755F10.1 | - | NA | | ENSG00000255320 | lncRNA | -1.6680829 | -0.3842963 | 8.70020766 | 0.00318174 | 0.43542912 |
| ENSG00000234500 | GS1-124K5.10 | K03121 | NA | | ENSG00000234500 | unprocessed_pseudogene | -0.9128438 | -0.3655969 | 8.68528377 | 0.0032079 | 0.43542912 |
| ENSG00000270130 | RP11-214K3.23 | - | NA | | ENSG00000270130 | lncRNA | -0.5547594 | 0.79447858 | 8.65940702 | 0.00325377 | 0.43542912 |
| ENSG00000258485 | SRMP2 | K00797 | NA | | ENSG00000258485 | processed_pseudogene | -0.9512417 | -0.5645284 | 8.65477867 | 0.00326205 | 0.43542912 |
| ENSG00000261574 | RP1-168P16.2 | - | NA | | NA | NA | -1.1349624 | 1.4911027 | 8.63871187 | 0.00329094 | 0.43542912 |
| ENSG00000247473 | CARS-AS1 | - | NA | | ENSG00000247473 | lncRNA | -0.5126052 | 0.60729991 | 8.62761217 | 0.00331105 | 0.43542912 |
| ENSG00000230581 | RP11-390F4.8 | K05692 | NA | | ENSG00000230581 | processed_pseudogene | -0.6422787 | 0.31945708 | 8.62044233 | 0.00332411 | 0.43542912 |
| ENSG00000168497 | SDPR | K19387 | | 8436 | ENSG00000168497 | protein_coding | 0.49879442 | 7.23502216 | 8.61918539 | 0.00332641 | 0.43542912 |
| ENSG00000250159 | RP11-381K20.2 | - | NA | | ENSG00000250159 | lncRNA | -0.5343155 | 0.5247808 | 8.58220618 | 0.00339464 | 0.43542912 |
| ENSG00000229728 | RP11-314N13.3 | - | NA | | ENSG00000229728 | lncRNA | -0.5247487 | 0.8405925 | 8.57838776 | 0.00340176 | 0.43542912 |
| ENSG00000245534 | RP11-219B17.1 | - | NA | | ENSG00000245534 | lncRNA | -0.4744596 | 2.65463068 | 8.55909456 | 0.003438 | 0.43542912 |
| ENSG00000236576 | RP11-22B10.3 | - | NA | | ENSG00000236576 | processed_pseudogene | -0.7223543 | 1.02486773 | 8.53033172 | 0.00349276 | 0.43542912 |
| ENSG00000244229 | RPL26P35 | K02898 | NA | | ENSG00000244229 | processed_pseudogene | -0.6495786 | -0.4590881 | 8.50185917 | 0.00354784 | 0.43542912 |
| ENSG00000119698 | PPP4R4 | K15426 | | 57718 | ENSG00000119698 | protein_coding | 0.6224526 | 0.11776759 | 8.47790367 | 0.00359486 | 0.43542912 |
| ENSG00000225300 | RP11-439E19.1 | - | NA | | ENSG00000225300 | lncRNA | -0.7284191 | 1.2147235 | 8.4591367 | 0.00363214 | 0.43542912 |
| ENSG00000250903 | GMDS-AS1 | - | NA | | ENSG00000250903 | lncRNA | -0.3849423 | 4.2375539 | 8.44486094 | 0.00366076 | 0.43542912 |

| | | | | | | | | | | | |
|---|---|---|---|---|---|---|---|---|---|---|---|
| ENSG00000178538 | CA8 | K01672 | | 767 | ENSG00000178538 | protein_coding | 0.58018021 | 1.72011573 | 8.44411366 | 0.00366227 | 0.43542912 |
| ENSG00000226705 | SDCBPP1 | K17254 | NA | | ENSG00000226705 | processed_pseudogene | -1.0848284 | -0.1764493 | 8.41923382 | 0.00371272 | 0.43542912 |
| ENSG00000243637 | AC019221.4 | - | NA | | NA | NA | -0.706938 | -0.1086778 | 8.41839186 | 0.00371444 | 0.43542912 |
| ENSG00000272498 | RP11-415F23.3 | - | NA | | ENSG00000272498 | lncRNA | -0.5427869 | 2.80750313 | 8.40167931 | 0.00374875 | 0.43542912 |
| ENSG00000230124 | RP5-1180C10.2 | - | | 84320 | ENSG00000230124 | protein_coding | -0.6910051 | 1.97814773 | 8.39954306 | 0.00375315 | 0.43542912 |
| ENSG00000122025 | FLT3 | K05092 | | 2322 | ENSG00000122025 | protein_coding | 0.59233982 | 3.12378094 | 8.37554554 | 0.00380303 | 0.43542912 |
| ENSG00000267694 | RP11-691H4.4 | - | NA | | ENSG00000267694 | lncRNA | -0.765662 | 0.72664129 | 8.37464987 | 0.00380491 | 0.43542912 |
| ENSG00000261451 | RP11-981G7.1 | - | NA | | ENSG00000261451 | lncRNA | -0.5503137 | 1.75711406 | 8.3722053 | 0.00381003 | 0.43542912 |
| ENSG00000260711 | RP11-747H7.3 | - | NA | | ENSG00000260711 | lncRNA | -0.6416419 | 3.2541186 | 8.35733955 | 0.00384133 | 0.43542912 |
| ENSG00000188536 | HBA2 | K13822 | | 3040 | ENSG00000188536 | protein_coding | 2.46026789 | 8.58469488 | 8.32661597 | 0.00390684 | 0.43542912 |
| ENSG00000264630 | RP11-4F22.2 | - | NA | | ENSG00000264630 | lncRNA | -1.164038 | 1.70746362 | 8.32258751 | 0.00391551 | 0.43542912 |
| ENSG00000185899 | TAS2R60 | K08474 | | 338398 | ENSG00000185899 | protein_coding | -0.9513838 | -0.3185196 | 8.2937732 | 0.00397812 | 0.43542912 |
| ENSG00000254254 | RP11-17A4.2 | - | NA | | ENSG00000254254 | lncRNA | -0.5536185 | -0.1655095 | 8.28134963 | 0.00400543 | 0.43542912 |
| ENSG00000264462 | MIR3648 | - | NA | | ENSG00000264462 | miRNA | -2.4334551 | 4.84918522 | 8.26822478 | 0.00403449 | 0.43542912 |
| ENSG00000227189 | AC092535.3 | - | NA | | ENSG00000227189 | lncRNA | -1.1436806 | -0.456219 | 8.26575249 | 0.00403999 | 0.43542912 |
| ENSG00000242716 | RNA5-8S5 | - | NA | | NA | NA | -1.307075 | 1.89357817 | 8.22893622 | 0.00412277 | 0.43542912 |
| ENSG00000235398 | LINC00623 | - | NA | | NA | NA | 0.48280967 | -0.0651094 | 8.22517215 | 0.00413133 | 0.43542912 |
| ENSG00000261308 | RP11-923I11.7 | - | | 401720 | ENSG00000261308 | protein_coding | -0.4544419 | 0.92082726 | 8.21613505 | 0.00415195 | 0.43542912 |
| ENSG00000169439 | SDC2 | K16336 | | 6383 | ENSG00000169439 | protein_coding | 0.56513838 | 0.65496335 | 8.20103147 | 0.00418666 | 0.43542912 |
| ENSG00000240356 | RPL23AP7 | K02893 | NA | | ENSG00000240356 | transcribed_processed_pseu | 0.53256584 | 2.75263848 | 8.18689088 | 0.00421942 | 0.43542912 |
| ENSG00000254879 | AC103828.1 | - | NA | | ENSG00000254879 | lncRNA | -0.8749204 | 1.26048295 | 8.1857995 | 0.00422196 | 0.43542912 |
| ENSG00000170011 | MYRIP | - | | 25924 | ENSG00000170011 | protein_coding | -0.5560325 | 1.66232411 | 8.18453199 | 0.00422491 | 0.43542912 |
| ENSG00000102554 | KLF5 | K09206 | | 688 | ENSG00000102554 | protein_coding | 0.49237617 | 1.96774472 | 8.17622084 | 0.00424431 | 0.43542912 |
| ENSG00000229052 | RP11-386I23.1 | K17985 | NA | | ENSG00000229052 | transcribed_processed_pseu | -0.7306023 | 1.64187799 | 8.16790808 | 0.00426381 | 0.43542912 |
| ENSG00000207129 | RNA5SP187 | - | NA | | ENSG00000207129 | rRNA_pseudogene | -0.4863946 | 0.63316606 | 8.16262206 | 0.00427625 | 0.43542912 |
| ENSG00000267702 | RP11-53B2.2 | - | NA | | ENSG00000267702 | lncRNA | -0.6823952 | 4.10014604 | 8.15986722 | 0.00428275 | 0.43542912 |
| ENSG00000184825 | HIST1H2AH | K11251 | NA | | NA | NA | -0.5869907 | 2.80966761 | 8.15764436 | 0.004288 | 0.43542912 |
| ENSG00000240935 | PLGLA | K01315 | NA | | ENSG00000240935 | transcribed_unprocessed_ps | -0.8201849 | -0.5013883 | 8.1532354 | 0.00429844 | 0.43542912 |
| ENSG00000244180 | AL592188.2 | - | NA | | NA | NA | -1.8235646 | 4.69288952 | 8.15232181 | 0.00430061 | 0.43542912 |
| ENSG00000100678 | SLC8A3 | K05849 | | 6547 | ENSG00000100678 | protein_coding | 0.64520648 | 0.25098556 | 8.14678766 | 0.00431375 | 0.43542912 |
| ENSG00000232564 | RP4-591N18.2 | - | NA | | ENSG00000232564 | lncRNA | -0.7202825 | 1.48679691 | 8.14370444 | 0.00432109 | 0.43542912 |
| ENSG00000128536 | CDHR3 | K16503;K114 | | 222256 | ENSG00000128536 | protein_coding | -0.5386647 | 2.39040826 | 8.141044 | 0.00432744 | 0.43542912 |
| ENSG00000196312 | HIATL2 | - | | 84278 | ENSG00000196312 | transcribed_unprocessed_ps | -0.3646159 | 2.9859268 | 8.13609868 | 0.00433925 | 0.43542912 |
| ENSG00000262074 | SNORD3B-2 | - | NA | | ENSG00000262074 | snoRNA | -1.1217256 | 0.40683584 | 8.12100494 | 0.00437553 | 0.43542912 |
| ENSG00000226803 | RP11-203B9.4 | - | NA | | ENSG00000226803 | lncRNA | -0.4090472 | 2.54123601 | 8.10966933 | 0.00440298 | 0.43542912 |
| ENSG00000244734 | HBB | K13823 | | 3043 | ENSG00000244734 | protein_coding | 3.30244583 | 9.6144834 | 8.10308024 | 0.00441901 | 0.43542912 |
| ENSG00000105982 | RNF32 | - | | 140545 | ENSG00000105982 | protein_coding | -0.4066835 | 1.76876839 | 8.09402861 | 0.00444113 | 0.43542912 |
| ENSG00000200397 | Y_RNA | - | NA | | ENSG00000200397 | misc_RNA | -0.4431899 | 0.39675556 | 8.0743391 | 0.00448965 | 0.43542912 |
| ENSG00000254481 | PTP4A2P2 | K18041 | NA | | ENSG00000254481 | processed_pseudogene | 0.50255239 | 0.90021554 | 8.06518284 | 0.00451239 | 0.43542912 |
| ENSG00000268367 | AC126614.1 | K02927 | NA | | NA | NA | -0.9876112 | -0.5321461 | 8.06411322 | 0.00451506 | 0.43542912 |
| ENSG00000228492 | RAB11FIP1P1 | K12484 | NA | | ENSG00000228492 | processed_pseudogene | -0.5886651 | 2.45479362 | 8.05718896 | 0.00453234 | 0.43542912 |
| ENSG00000223899 | SEC13P1 | K14004 | NA | | ENSG00000223899 | processed_pseudogene | -0.7329607 | -0.3933527 | 8.05228604 | 0.00454463 | 0.43542912 |
| ENSG00000266378 | RP11-214O1.3 | - | NA | | ENSG00000266378 | lncRNA | -1.0299226 | 0.80746186 | 8.04219452 | 0.00457001 | 0.43542912 |

Mass cytometry panel used for immunophenotyping

| **Metal Isotope /Fluorochrome** | **Antigen** | **Clone** | **Manufacturer** |
|---|---|---|---|
| Barcoding and fluorescent | | | |
| $^{104}Pd$, $^{106}Pd$ or $^{108}Pd$ | CD45 | 30-F11 | BD/Biolegend |
| Biotin | CD86 | IT2.2 | BD |
| AF647 | CD160 | BY55 | BD |
| Remaining surface stain | | | |
| $^{89}Y$ | CD11c | Bu15 | Biolegend |
| $^{113}In$ | IgM | G20127 | BD |
| $^{115}In$ | Biotin | 1D4-C5 | Biolegend |
| $^{139}La$ | CD56 | NCAM16.2 | BD |
| $^{141}Pr$ | CD27 | M-T271 | BD |
| $^{142}Nd$ | CD19 | HIB19 | Biolegend |
| $^{143}Nd$ | CD45RA | HI100 | BD |
| $^{144}Nd$ | TCRgd | B1 | Biolegend |
| $^{145}Nd$ | CD4 | RPA-T4 | Biolegend |
| $^{146}Nd$ | IgD | IA6-2 | BD |
| $^{147}Sm$ | Cy5 (for AF647) | CY5-15 | Sigma |
| $^{149}Sm$ | CD366 (Tim3) | 7D3 | BD |
| $^{150}Nd$ | KLRG1 | SA231A | Biolegend |
| $^{151}Eu$ | CD123 | 6H6 | Biolegend |
| $^{152}Sm$ | CD45RO | UCHL1 | Biolegend |
| $^{153}Eu$ | CD62L | DREG | Biolegend |
| $^{154}Gd$ | CD3 | UCHT1 | BD |
| $^{156}Gd$ | CD279 (PD-1) | EH12 .2H7 | Biolegend |
| $^{158}Gd$ | CD87 | VIM5 | Biolegend |
| $^{159}Tb$ | CD223 (Lag3) | 17B4 | Genetex |
| $^{161}Dy$ | CD274 (PD-L1) | MIH1 | BD |
| $^{165}Ho$ | CD16 | 3G8 | BD |
| $^{166}Er$ | TIGIT | MBSA43 | eBioscience |
| $^{167}Er$ | CD66a | YTH71.3 | Abcam |
| $^{168}Er$ | CD8a | RPA-T8 | BD |
| $^{169}Tm$ | CD25 | M-A251 | Biolegend |
| $^{170}Er$ | CD152 (CTLA4) | 14D3 | eBioscience |
| $^{172}Yb$ | CD197 (CCR7) | G043H7 | Biolegend |

| **Metal Isotope /Fluorochrome** | **Antigen** | **Clone** | **Manufacturer** |
|---|---|---|---|
| Intracellular stain | | | |
| $^{148}Nd$ | Helios | 22F6 | Nobus Bio |
| $^{155}Gd$ | EOMES | WD1928 | eBioscience |
| $^{162}Er$ | FOXP3 | PCH101 | eBioscience |
| $^{163}Dy$ | BLIMP | 6D3 | BD |
| $^{164}Er$ | GATA3 | L50-823 | BD |
| $^{171}Yb$ | GranzymeB | REA226 | Miltenyi |
| $^{175}Lu$ | Perforin | dG9 | Biolegend |
| $^{191/193}Ir$ | DNA Intercalator | - | Fluidigm |
| $^{209}Bi$ | T-bet | 4B10 | BD |
